# Real World Predictors of Response and 24-month survival in high-grade *TP53*-mutated Myeloid Neoplasms

**DOI:** 10.1101/2024.03.11.24303995

**Authors:** Amandeep Kaur, Alexandra E. Rojek, Emily Symes, Anand A. Patel, Mariam Nawas, Jay L. Patel, Payal Sojitra, Barina Aqil, Madina Sukhanova, Megan E. McNerney, Leo Wu, Aibek Akmatbekov, Jeremy Segal, Melissa Tjota, Sandeep Gurbuxani, Jason X. Cheng, Su-Yeon Yeon, Harini V. Ravisankar, Carrie Fitzpatrick, Angela Lager, Michael W. Drazer, Caner Saygin, Pankhuri Wanjari, Panagiotis Katsonis, Olivier Lichtarge, Jane E. Churpek, Sharmila B. Ghosh, Ami B. Patel, Madhu P. Menon, Daniel A. Arber, Peng Wang, Girish Venkataraman

## Abstract

Current therapies for high-grade *TP53*-mutated myeloid neoplasms (≥ 10% blasts) do not offer a meaningful survival benefit except allogeneic stem cell transplantation in the minority who achieve a complete response to first line therapy (CR1). To identify reliable pre-therapy predictors of complete response to first-line therapy (CR1) and outcomes, we assembled a cohort of 238 individuals with *TP53*-mutated myeloid neoplasms and ≥ 10% blasts with well-annotated clinical, molecular and pathology data. Key outcomes examined were CR1 & 24-month survival (OS24). In this elderly cohort (median age 68.1 years) with 74.0% receiving frontline non-intensive regimens (hypomethylating agents+/-venetoclax), the overall cohort CR1 rate was 26.4% (51/193). We additionally identified several pre-therapy factors predictive of inferior CR1 including male gender (*P =* .015), ≥ 2 autosomal monosomies (P < .001), -17/17p (*P =* .015), multi-hit *TP53* allelic state (*P* < .001) and *CUX1* co-alterations (*P =* .011). In univariable analysis of the entire cohort, inferior OS24 was predicated by ≥ 2 monosomies (*P =* .004), *TP53* VAF>25% (P < .001), *TP53* splice junction mutations (*P =* .008) and antecedent treated myeloid neoplasm (*P =* .001). In addition, mutations/deletions in *CUX1*, *U2AF1*, *EZH2*, *TET2*, *CBL*, or *KRAS* (’*EPI6*’ signature) predicted inferior OS24 (HR = 2.0 [1.4–2.7]; *P <* .0001). In a subgroup analysis of HMA +/-Ven treated individuals (*N =* 141), *TP53* VAF and monosomies did not impact OS24. A risk score for HMA +/-Ven treated individuals incorporating three pre-therapy predictors including *TP53* splice junction mutations, *EPI6* and antecedent treated myeloid neoplasm stratified 3 prognostic distinct groups: intermediate, intermediate-poor, and poor with significantly different median (12.6, 6.3, 4.3 months) and 24-month (19.3%, 5.2%, 0.5%) survival (P < .001). For the first time, in a seemingly monolithic high-risk cohort, our data identifies several baseline factors that predict response and 24-month survival.

**Key Points:**

- ≥2 monosomies, multi-hit *TP53* allelic state & *CUX1* deletions are associated with poor frontline response in *TP53*-mutated myeloid neoplasm.
- *TP53* splice junction mutations and co-alterations in any one of six genes (*CBL, CUX1, EZH2, KRAS, TET2 & U2AF1*) adversely impact 24-month survival.

## INTRODUCTION

Several studies in the past decade have confirmed the adverse outcome of complex and monosomal karyotype as well as *TP53* alterations in the context of myeloid neoplasms (MN).^1–4^ *TP53^MUT^* MN are characterized by frequent complex karyotype (CK) and very poor 2-year survival of 12.8%, regardless of blast count.^5^ Based on these studies, high-risk myelodysplastic syndrome (MDS) including MDS/AML (MDS/AML) and AML with mutated *TP53* (*TP53^MUT^*) are now recognized as distinct categories in the World Health Organization 5^th^ edition (WHO5), International Consensus Classification of hematopoietic neoplasms (ICC), and European LeukemiaNet (ELN) risk stratification.^6–9^

Beyond assessing complete response to first-line therapy (CR1) and allogeneic hematopoietic stem cell transplantation (allo-SCT) in CR1, there are no well-established pre-therapy prognostic indicators in this cohort of patients^10^ who are automatically assigned to the adverse group per ELN2022 risk stratification.^9^ Furthermore, there is a significant heterogeneity in responses to available therapies (intensive chemotherapy *vs.* hypomethylating agent (HMA)-based therapy) or agents used in clinical trials (for e.g. APR-246^11^) with few long-term survivors even in those receiving allo-SCT.^12^ Consequently, there is a pressing need to explore better means to identify disease characteristics influencing therapeutic response to optimally select frontline treatments and identify patients who are likely to achieve durable benefit from allo-SCT in CR1.

With this background, in this study we asked if 1) specific chromosomal alterations within a CK (such as autosomal monosomies), 2) the type of *TP53* mutation (missense [*TP53^MIS^*] *vs.* non-missense *TP53* mutations [*TP53^NMIS^*]), and 3) patterns of co-mutations/alterations contribute additional prognostic value beyond *TP53* allelic state in risk-stratifying *TP53^MUT^* MN.

## MATERIALS AND METHODS

### Cohort Case Selection and Sample Procurement

We identified patients with *TP53^MUT^* MN carrying ≥ 1 *TP53* mutation at a VAF ≥ 3% diagnosed between 2014-2023 across four US centers with largely similar treatment practices. We excluded individuals with any of the following: a known germline *TP53* mutation, known *TP53^MUT^*precursor states (CHIP, CCUS), MDS with mutated *TP53* (< 10% blasts) ^6^, and core binding factor-altered AMLs.

#### Data Collection

We collected data on demographics, marrow pathology, molecular and cytogenetic information, and treatment types. Therapies were categorized as: **1]** Intensive Chemotherapy (IC) (7+3 or high-dose cytarabine), **2]** Hypomethylating agent (HMA)-based (without venetoclax), **3]** HMA-based with venetoclax, or **4]** Best supportive care/palliative regimens. Response was assessed per ELN 2017 guidelines^13^ denoting both CR and CR with incomplete hematologic recovery (CRi) as a composite measure of CR1.

### Cytogenetic studies

Chromosome analysis was performed following standard cytogenetic laboratory clinical protocol. Fluorescence in situ hybridization (FISH) testing was performed using probe sets targeting most recurring abnormalities in myeloid neoplasms on bone marrow or malignancy-involved peripheral blood. Using previously published criteria, over 90% of the cohort cases met the classification of monosomal karyotype (MK).^1^ Therefore, we applied a revised definition of MK restricting to only unique autosomal monosomies (0-1 *vs.* 2+) without considering other structural alterations (**supplemental S1**).

### Next Generation Sequencing

Somatic next-generation sequencing (NGS) data was available in all cases. To maintain consistency in our analysis, we only included genes that were tested across two or more centers (**supplemental S2**). For missense *TP53* mutations, we also examined the evolutionary action score for p53 (**supplemental S3**).^14^ Additionally, germline testing data (**supplemental S4**) was available in a subset of cases.

#### Allelic status

*TP53* multi-hit (*TP53^MH^*) allelic state was designated per ICC 2022 schema.^6^ However, single-hit (*TP53^SH^*) designation used an expanded VAF cutoff including cases with VAF between 3-10%. Additionally, we modeled any non-missense *TP53* mutations (*TP53^NMIS^*) as a binary predictor.

### Data analysis

The primary endpoint was 24-month overall survival (OS24) from the time of diagnosis of *TP53^MUT^* MN to death or last follow-up, censoring patients alive at 24 months. Response to first-line therapy was also evaluated. We employed non-parametric Kaplan-Meier methods, along with Cox proportional hazards (P-H) regression models^15^, and, where suitable, flexible parametric models^16^ (**supplemental S5**).

## RESULTS

### Cohort Summary

Table 1 depicts the baseline data of all patients with therapy/response data of all patients receiving definitive therapy stratified by *TP53* allelic state at diagnosis of *TP53^MUT^* myeloid neoplasm. Individuals accrued after 2018 were significantly older with 47.7% surpassing 70 years of age as opposed to 30.6% in the pre-2018 period (*P =* .020). Additionally, post-2018 individuals frequently received lower-intensity therapies including HMA+/-venetoclax or CPX-351. Furthermore, individuals with AML (blasts ≥ 20%) were significantly more likely to receive VEN-based regimens compared to patients with MDS/AMLs (*P =* .003) at diagnosis. Most patients (80.4%; 172/214) exhibited complex karyotypes characterized by frequent autosomal monosomies (83.6%; 179/214) along with several recurrent balanced and unbalanced structural alterations in 93.5% of cases (**Figure 1A**-1C ). The two most prevalent single autosomal monosomies affected chromosomes 17 in 32.7% (70/214) and 7 in 30.8% (66/214) (**Figure 1B** for co-occurrence plot). There was no association between *TP53* allelic state and prior cytotoxic-therapy (*P =* .15). Among germline alterations, *BRCA1*, *BRCA2* and *DDX41* alterations predominated.

**Figure 1:**
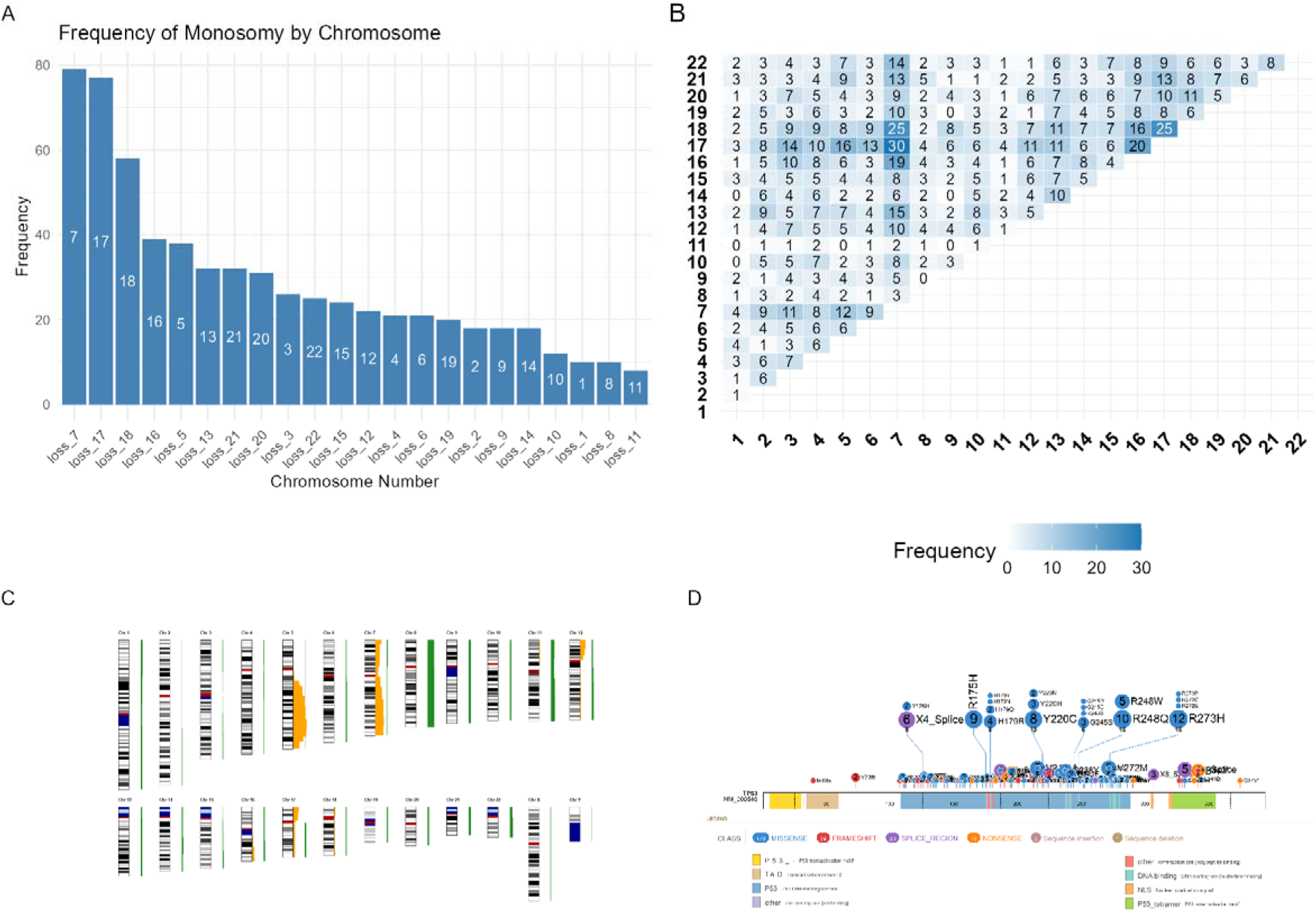
Summary of cytogenetic alterations and *TP53* mutations. **A.** Frequencies of autosomal monosomies (percentages represent frequencies in patients with available data on karyotype). The most frequent autosomies involved chromosomes 7, 17 and 18. **B.** Co-occurrence matrix of autosomal monosomies. Counts indicate frequencies of co-occurrence. The most frequently co-occurring monosomies with loss of 17 were monosomies 7 and 16, followed by monosomies of chromosomes 5, 6 and 12. **C.** Ideogram of global losses (orange) and gains (green) across the genome. Recurrent losses are enriched in chromosomes 5, 7 and 17. Furthermore, we observed additional recurrent losses on 12p, besides 16q, 18p, and 18q. Chromosomal gains were prevalent, particularly on chromosomes 8 (trisomy 8), as well as on chromosomes 1, 9, 11, 21, and 22. Clean karyotypes were batched parsed in CytoGPS to create .json files with loss, gain & fusion information. These files were examined using the RCytoGPS package for R, treating them as a binary matrix, to create the ideogram.^35^. **D.** Summary of *TP53* mutations in the cohort. Somatic variants in *TP53* visualized using lollipop plot generated via the ProteinPaint web-based application.^36^ Majority comprised hot-spot DNA-binding domain missense mutations. Chromosomal position coordinates were culled from the IARC database of *TP53* mutations. A few complex mutations seen in a few cases are not depicted and numbers may not match up with that depicted in the results section.

**Table 1:** Baseline characteristics of all 238 patients at diagnosis of *TP53^MUT^* Myeloid Neoplasm stratified by *TP53* allelic state at diagnosis (see methods section for definition of allelic state).

|  | *TP53*<sup>SH</sup><br>N=67 (28.2%) | *TP53*<sup>MH</sup><br>N=171 (71.8%) | Test |
| --- | --- | --- | --- |
| **Age at diagnosis, Median [IQR]** |  |  |  |
| Age (years) | 68.3 [14-89] | 67.9 [23-93] | 0.597 |
| **Baseline labs, Median [IQR]** |  |  |  |
| Hemoglobin (g/dL) | 8.2 [3-13] | 8.0 [4-17] | 0.826 |
| Platelet Count (10<sup>9</sup>/L) | 51.0 [5-344] | 40.0 [5-585] | 0.711 |
| Abs. Neutrophil Count (10<sup>9</sup>/L) | 0.6 [0-5200] | 0.7 [0-8740] | 0.571 |
| **Sex** |  |  |  |
| Male | 32 (47.8%) | 102 (59.6%) | 0.096 |
| Female | 35 (52.2%) | 69 (40.4%) |  |
| **Era of dx.-no.(%)** |  |  |  |
| Pre-2018 | 19 (28.4%) | 43 (25.1%) | 0.612 |
| 2018-current | 48 (71.6%) | 128 (74.9%) |  |
| **Prior Treated Myeloid Neoplasm\*-no.(%)** |  |  |  |
| No | 53 (79.1%) | 138 (80.7%) | 0.781 |
| Yes | 14 (20.9%) | 33 (19.3%) |  |
| **MDS/AML-ICC 2022** |  |  |  |
| MDS/AML (10-19% blasts) | 11 (16.7%) | 35 (20.5%) | 0.507 |
| AML (20%+ blasts) | 55 (83.3%) | 136 (79.5%) |  |
| **Complex karyotype** |  |  |  |
| Absent | 21 (37.5%) | 21 (13.3%) | <0.001 |
| Present | 35 (62.5%) | 137 (86.7%) |  |
| **Chromosome 7 loss** |  |  |  |
| Absent | 31 (54.4%) | 73 (45.3%) | 0.240 |
| Present | 26 (45.6%) | 88 (54.7%) |  |
| **Monosomies-no.(%)** |  |  |  |
| 0-1 Monosomy | 32 (57.1%) | 49 (31.0%) | <0.001 |
| 2+ Monosomies | 24 (42.9%) | 109 (69.0%) |  |
| **Chromosome 12(p) loss** |  |  |  |
| del(12p) absent | 63 (94.0%) | 141 (82.5%) | 0.022 |
| del(12p) present | 4 (6.0%) | 30 (17.5%) |  |
| ***TP53* mutation class†-no.(%)** |  |  |  |
| Missense | 48 (71.6%) | 108 (63.2%) | 0.215 |
| Non-Missense | 19 (28.4%) | 63 (36.8%) |  |
| **Germline alteration.-no.(%)** |  |  |  |
| Absent | 20 (76.9%) | 46 (86.8%) | 0.266 |
| Present | 6 (23.1%) | 7 (13.2%) |  |
| **Co-alterations-no.(%)** |  |  |  |
| Absent | 29 (43.3%) | 61 (35.7%) | 0.276 |
| Present | 38 (56.7%) | 110 (64.3%) |  |
| **Therapies-no.(%)** |  |  |  |
| Low-inten. (HMA±VEN) | 41 (64.1%) | 104 (62.7%) | 0.817 |
| Intensive (Chemo) | 14 (21.9%) | 37 (22.3%) |  |
| Best.Supp.Care | 7 (10.9%) | 15 (9.0%) |  |
| CPX-351/Vyxeos | 2 (3.1%) | 10 (6.0%) |  |
| **Resp. frontline†-no.(%)** |  |  |  |
| No CR/CRi | 31 (56.4%) | 111 (80.4%) | <0.001 |
| CR/CRi | 24 (43.6%) | 27 (19.6%) |  |

Med., Median; IQR, Interquartile range; BSC, Best Supportive Care; CR, Complete Response; CRi, CR & incomplete hematologic recovery
\*These included treated or untreated MDS (Low and High-risk), MDS/MPN and MPN without any *TP53*<sup>MUT</sup> up until evolution/progression to a *TP53*<sup>MUT</sup> myeloid neoplasm.
†Numbers reported in therapies and response may not add up to cohort total since some patients were either untreated or if treated, response could not be evaluated due to various reasons (active second malignancy, early treatment-emergent adverse effects, transferred care elsewhere or early mortality)
‡In pts with multiple mutations, case was designated *TP53*<sup>NMIS</sup> if ≥1 *TP53* mutation was a *TP53*<sup>NMIS</sup> mutation.

### *TP53^NMIS^* mutations are frequently associated with multi-hit allelic state and autosomal monosomies

Mutational analysis identified 300 pathogenic mutations (196 unique mutations) among the 240 patients comprising mostly hot-spot DNA-binding domain *TP53^MIS^* mutations (**Figure 1D**). A total of 30.0% (72/240) harbored non-missense *TP53* mutations, either singly or as part of multiple *TP53* mutations. Individuals with single *TP53* mutations were significantly more likely to harbor loss of chromosome 17p either due to monosomy 17 or structural losses of 17p (45.3% *vs.* 27.3% in multiple *TP53* mutations; *P =* .019). Among *TP53^NMIS^* mutations, splice junction mutations (*TP53^sjm^*) occurring in 22 (9.2%) were significantly more prevalent in men (*P =* .041).

The median *TP53* VAF was 42% (IQR: 23–59.69%). and did not differ between *TP53^MIS^* and *TP53^NMIS^* alterations (*P =* .73). Sub-clonal *TP53* mutations (VAF < 10%) were observed in 6.7% (16/240) with single mutations and 4.2% (10/240) with 1+ mutations. *TP53^NMIS^*were more frequently associated with ≥ 2 monosomies (74.0% *vs.* 56.0% in *TP53^MIS^*; *P =* .010), chromosome 7 losses (66.2% *vs.* 45.1% in *TP53^MIS^*; *P =* .003), multiple *TP53* mutations (41.5% *vs.* 15.4% in *TP53^MIS^*; *P <* .0001) and germline alterations (33.3% *vs.* 10.9% with germline in *TP53^MIS^*; *P =* .022). Patients with *TP53^MIS^*EAp53 score in upper quartile (*N =* 101/138) frequently had *TP53* VAF > 25% (87.0% *vs.* 73.2%*; P =* .06).

### A 6-gene co-alteration signature including *CUX1* deletion is associated with higher *TP53* VAF

Co-alterations were present in 62.2% (*N =* 148) with a median of 1 co-alteration (range: 0–16 co-alteration). Co-alterations in epigenetic pathways (*DNMT3A* and *TET2*) genes predominated (**Figure 2**), in line with prior observations.^5^ Males had a higher frequency of mutations in genes of the spliceosome complex (*SF3B1*, *SRSF2*, *U2AF1* or *ZRSR2*) (19.4% *vs.* 5.8% in females; *P =* .002) and the nine myelodysplasia-related genes^6^ (32.1% *vs.* 19.2% in females; *P =* .026). Among structural alterations detected by NGS, losses at the *CUX1* locus occurring in 9.9% (22/223) of individuals were the most frequent structural alteration (after losses at the *TP53* locus). Patients with any co-alteration had a significantly higher median *TP53* VAF (45.6% *vs.* 42.1%; *P =* .012). Likewise, individuals carrying *CUX1*-alterations had almost double the *TP53* VAF compared to those without *CUX1* alterations (78.5% *vs.* 43%; *P <* .0001).

**Figure 2:**
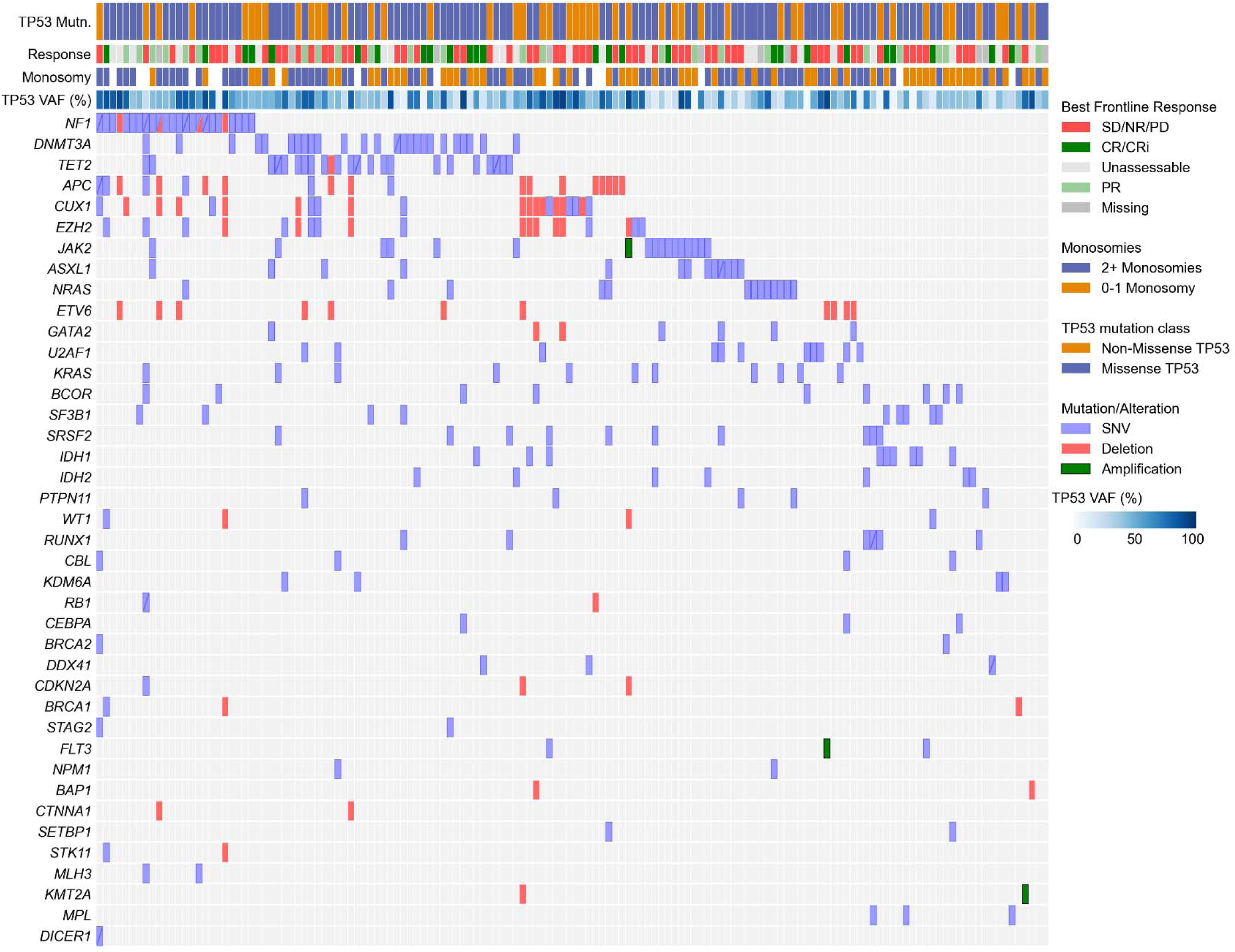
Co-mutations and structural alterations in patients with any co-mutations. Only variants occurring at least twice within the cohort are represented. Notably, structural alterations/losses were common with certain genes, particularly *CUX1*, *EZH2*, and *APC*.

We constructed an ’*EPI6*’ signature based on alterations co-occurring in ≥ 4 individuals and defined by the presence of mutations or deletions in at least one of six key genes associated with methylation (*TET2*, *EZH2*), mitogen-activated protein kinase (*KRAS*), signaling (*CBL*), splicing (*U2AF1*) pathways or *CUX1*. We selected these genes based on their frequency of occurrence and impact in initial exploratory univariable analysis. This *EPI6* signature was present in 25.6% (61/238) of individuals with a significantly higher proportion harboring *TP53* VAF > 25% (88.5% *vs.* 74.6% with VAF > 25% in individuals lacking *EPI6*; *P =* .023). We used this *EPI6* signature going forward in our analyses of treatment outcomes and long-term survival. Response data: *CUX1* alterations and ≥ 2 autosomal monosomies predict inferior CR1

#### First-line Treatment and Response Information

Among the 230 patients, low-intensity regimens (mostly HMA-based) were used in 145 (63.0%), intensive chemotherapy in 22.2% (*N =* 51), CPX-351 in 12 (5.2%) while 22 (9.6%) received only best supportive care. In a minority of patients, treatment information was not available (lost to follow up). Among HMA-based therapies (*N =* 141), HMA (with or without magrolimab) was used in 59 (41.8%) while HMA + VEN was used in 58.2% (*N =* 82). A total of 15.1% (36/238) went on to receive allogeneic stem cell transplantation with 72.4% receiving myeloablative conditioning regimens. First-line response was evaluable in 193 patients with 26.4% achieving CR, 22.8% with partial response, and 50.8% with non-response/stable-progressive disease. A small subset of patients could not be evaluated for treatment response because of treatment discontinuation due to treatment-emergent adverse effects, developed active second non-hematopoietic malignancies, or were lost to follow-up shortly after diagnosis. These patients were excluded from all response analyses but were included in the analysis of 24-month overall survival (OS24).

#### Univariable Predictors of inferior CR1

There was no difference in CR rates between lower-intensity (30.0%; *N =* 36/120) and intensive regimens (25.0%; *N =*12/48, *P =* .52). although surprisingly, CR rates were higher in females (35.4% *vs.* 19.8% CR1 in Male; *P =* .015). Looking at the entire cohort (intensively and non-intensively treated patients), the biological predictors of inferior CR1 were ≥ 2 monosomies (16.8% *vs.* 40.8% CR1 in 0-1 Monosomy; *P <* .001), losses on chromosome 17 (16.7% *vs.* 33.0%; *P =* .015), *TP53^MH^* allelic state (19.6% *vs.* 43.6%; *P <* .001), myelodysplasia-related gene mutations (15.0% *vs.* 31.6%; *P =* .016) as well as *EPI6* (16.3% *vs.* 29.9%; *P =* .06). Remarkably, none of the individuals with *CUX1* alterations achieved a CR (0.0%; 0/18) *vs.* 44/161 (27.3%) CR1 without *CUX1* alterations. Chromosome 5 (*P =* .56) and chromosome 7 losses (*P =* .38) did not impact frontline response. Among the intensively treated subgroup, hot-spot *TP53* mutations were associated significantly inferior frontline response (17.9% *vs.* 57.1% CR1 in Non-hotspot; *P =* .025).

Among individuals treated with HMA-based regimens, HMA+VEN did not result in significantly higher CR rates (34.3%) *vs.* 24.0% in HMA only (*P =* .23). These data are in line with several recent reports.^17–20^ However, among patients with blast counts over 20%, HMA+Ven resulted in marginally higher CR rates (34.5% *vs.* 16.7% in HMA only; *P =* .08).

#### Multivariable response prediction models in entire cohort and HMA subgroup

In a multi-variable logistic regression model on the entire cohort (*N =* 167) including gender and the aforementioned biologic predictors (monosomies, *TP53* allelic state, gender and *EPI6*), ≥ 2 monosomies (OR = 0.31 [95% CI: 0.15–0.67]; *P =* .003), and *TP53* allelic state (OR = 0.43 [95% CI: 0.20–0.95]; *P =* .037) predicted significantly inferior response with a marginal effect for *EPI6* (*P =* .09) (**Supplemental Figure 1**, ROC curve). Among the subgroup treated only with HMA-based therapies (*N =* 113), the logistic regression model identified ≥ 2 monosomies (OR = 0.24 [95% CI: 0.10–0.59]; *P =* .002), and *TP53* allelic state (OR = 0.37 [95% CI: 0.14–0.92]; *P =* .033) while *EPI6* (*P =* .52) was not relevant.

### Baseline outcome data

The median duration of follow up from diagnosis of *TP53^MUT^* MN to study exit was 6.3 months (range: 0.2–72.8 months) with 203 deaths in 238 patients and a death rate of 88.5 per 1000 patient-years with a 24-month survival of 15.3% (95% CI = 10.7–20.7%).

### Uni-variable analyses of OS24

Age at diagnosis > 70 years (HR = 1.5 [1.2–2.1]; *P =* .004), antecedent treated myeloid neoplasm (HR = 1.8 [1.2–2.5]; *P =* .002; **Figure 3A**) and complex karyotype (HR = 1.8 [1.1–2.7]; *P =* .009) all predicted worse OS24. Blast count at diagnosis and prior cytotoxic therapy-related myeloid neoplasm did not influence outcomes. Among chromosomal alterations, del(7q) (HR = 1.5 [1.1–2.1]; *P =* .007) and monosomy of chromosomes 17 (HR = 1.4 [1.0–1.9]; *P =* .050) predicted inferior outcomes. Neither del(5q), del(17p), nor monosomies of 5 or 7 impacted survival. Comparing 0-1 *vs.* ≥ 2 monosomies, the latter group experienced significantly higher hazard of mortality (HR = 1.7 [1.2–2.3]; *P =* .002) (**Figure 3B**).

**Figure 3:**
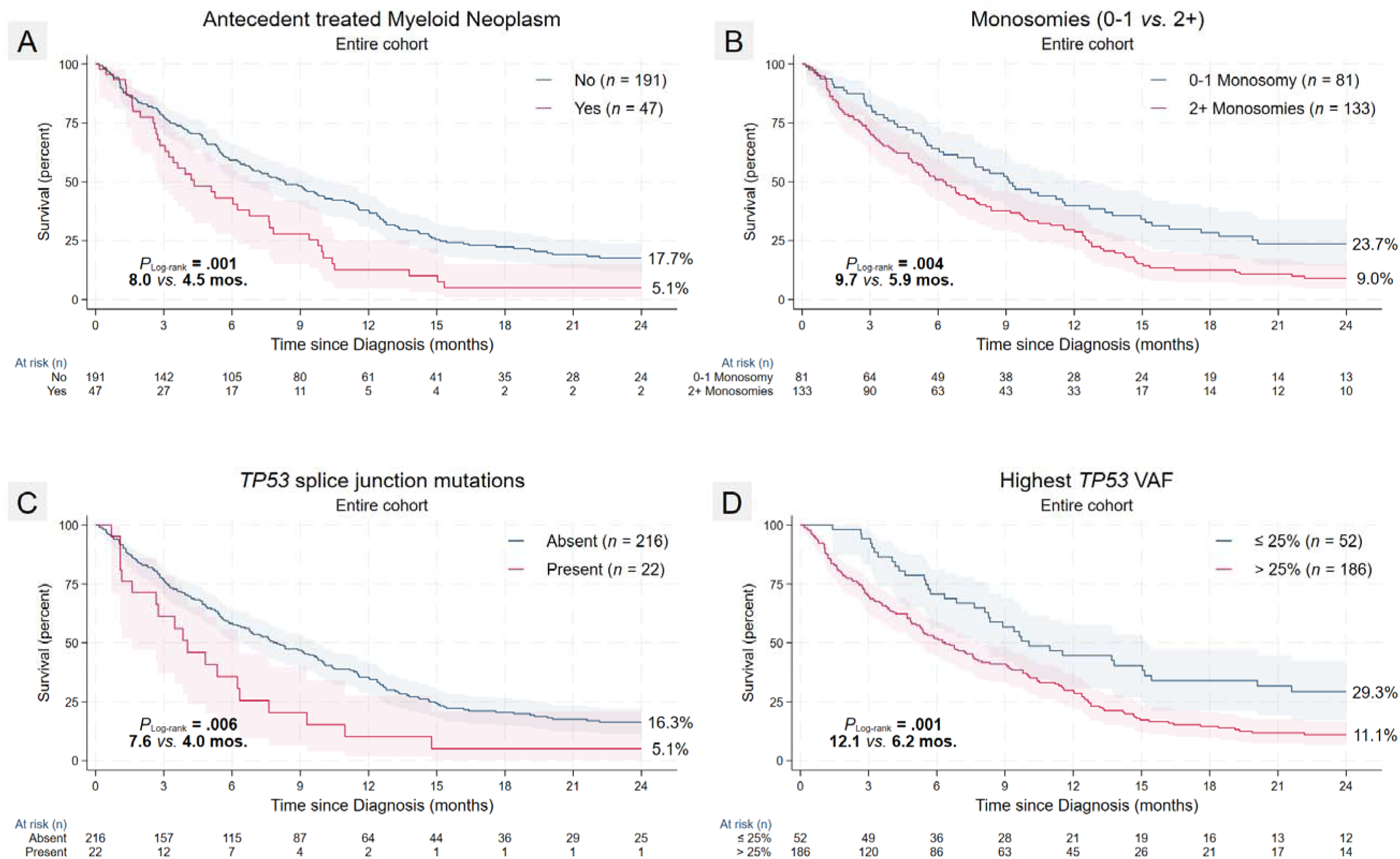
Univariable K-M plots of key pre-therapy adverse predictors. **A.** Antecedent myeloid neoplasm, **B.** Autosomal monosomies and **D.** Highest *TP53* VAF ≥ 25%, all adversely impact overall survival (OS24). **C.** *TP53* splice junction mutations were significantly associated with male gender and were associated with worse survival compared to all other classes of *TP53* mutations.

#### TP53^NMIS^ but not multi-hit allelic state is adverse

We next assessed different measures of *TP53* mutations and their associations with outcome. The number of *TP53* mutations (1 *vs.* 1+) did not impact OS24. Overall, *TP53^NMIS^* did not impact OS24, among individuals with single *TP53* mutations, *TP53^NMIS^* predicted significantly inferior median survival (4.0 *vs.* 9.1 mos.; *P_Log-rank_* = .032) with early mortality. Among *TP53^NMIS^*, splice junction mutations were associated with particularly poor outcomes (HR = 1.9 [1.2–3.1]; *P =* .007) (**Figure 3C**). Neither underlying germline alterations (*P =* .39) nor *TP53^MH^* allelic state (*P =* .12) conferred worse outcomes.

While all *TP53* VAF cutoffs (>10%, >25%, >40%, and >50%) predicted inferior OS24, we selected VAF > 25% (HR = 1.8 [1.3–2.7]; *P =* .001) for all subsequent analysis based on the balance of cases across both groups for an appropriately powered analysis (**Figure 3D**). Intriguingly, the beneficial impact of a lower VAF within each of these binary cut-points were restricted only to male gender when examined separately by gender (**Supplemental Figure 2**) although the analysis was slightly underpowered.

#### CUX1 deletions and EPI6 signature including CUX1 are both adverse

The presence of any co-alteration occurring in 148/238 (62.2%) did not impact survival (*P =* .60). *CUX1* alterations (mostly losses detected by NGS) were associated with particular very poor survival (1.1% *vs.* 15.0%; P*_fpm_* < .001) without any survivor beyond 12 months of diagnosis. Looking at combinations of co-altered genes, co-alterations in myelodysplasia-related genes (*P =* .62) or spliceosome complex genes did not impact survival (*P =* .84). In an age-adjusted model, *EPI6* predicted significantly inferior OS24 (3.3% *vs.* 17.9% for no *EPI6*; P*_fpm_* < .0001; HR = 2.0 [1.4– 2.7]; *P <* .0001 **Figure 4**) with a differential impact of each of *EPI6* genes when stratified by gender (**Figure 4A**).

**Figure 4:**
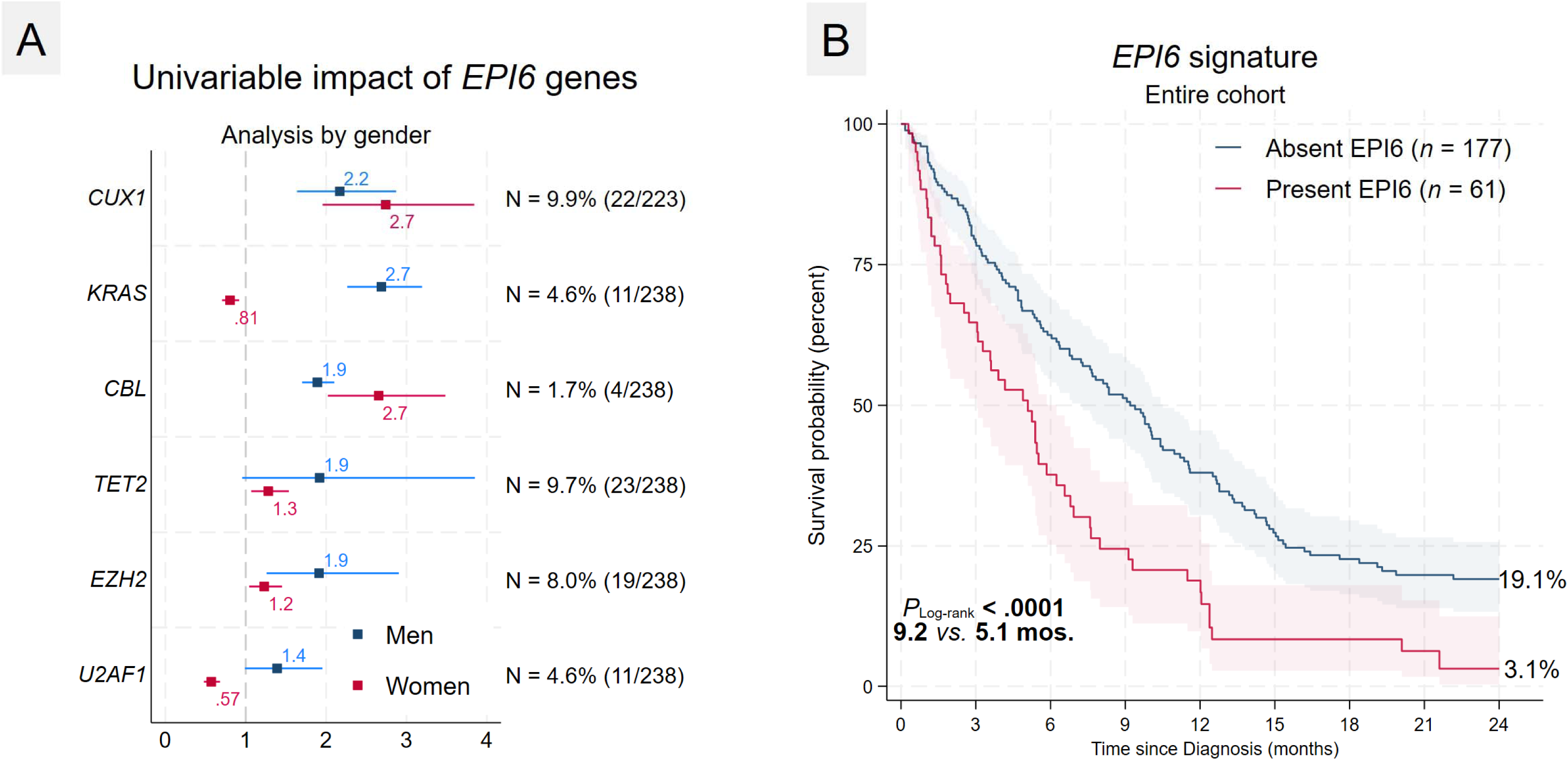
Univariable adverse impact of *EPI6* genes and outcome. **A.** Forest plot of the hazard ratios of each of the seven “*EPI6*” genes in the OS24 analysis with numbers and frequencies of occurrence depicted on the right. There were gender specific differences in some of the genes and hence these genes were included in the *EPI6* signature despite borderline adverse significance in the combined analysis. **B.** Patient carrying co-mutations/alterations in any of the 6 “*EPI6*” genes including *CBL*, *CUX1*, *EZH2*, *TET2*, *KRAS*, or *U2AF1* experienced shorter median and overall survival. *CUX1* was the most influential gene in the *EPI6* signature with overall survival of *CUX1*-altered individuals approaching 0% by 12 months after diagnosis.

#### Venetoclax-based regimens do not offer significant survival benefit

Using low-intensity therapies as a referent group, patients receiving intensive regimens (7+3 or HiDAC-based) experienced slightly better outcomes in an age-adjusted analysis (HR = 0.7 [0.5–1.0]; *P =* .07; *N =* 196) and OS24 (27.2% *vs.* 12.1% in low-intensity; *P_fpm_ =* .014). See **Supplemental Figure 3** for outcomes by major therapy classes. Among patients treated with intensive chemotherapy (excluding patients treated with CPX-351), VAF > 25% (HR = 3.3 [1.3–8.5]; *P =* .016; *N =* 51) and EAp53 score in the highest quartile (HR = 1.9 [0.9–4.2]; *P =* .11; *N =* 39) were both adverse with a marginal effect for latter. In a subgroup analysis by VAF, the beneficial effect of intensive regimens over non-intensive regimens was restricted only to individuals with VAF ≤ 25%.

Among HMA-treated individuals, HMA+venetoclax regimens were marginally adverse in patients older than 70 years (HR = 1.5 [0.9–2.6]; *P =* .15; *N =* 73) with no overall survival benefit compared to HMA only, in line with recent data.^21^ Within this subgroup (*N =* 141), only antecedent treated myeloid neoplasm (HR = 1.7 [1.1–2.8]; *P =* .020; *N =* 141), *TP53^sjm^* (HR = 2.0 [1.0–3.8]; *P =* .045; *N =* 141) and *EPI6* predicted poor outcomes (HR = 2.0 [1.0–3.8]; *P =* .045; *N =* 141). *TP53* allelic status, *TP53* VAF, monosomies and EAp53 score (**Supplemental Figure 4**) were not prognostic in this subgroup (**Supplemental Figure 5**). Achieving CR1 to first-line significantly improved outcomes (OS24 39.4% *vs.* 7.3% for no CR1; *P <* .0001) overall and. within treatment subgroups of intensively treated as well as HMA-treated individuals (OS24 31.4% *vs.* 3.4% for HMA+/-Ven subgroup; *P <* .0001).

#### Allogeneic transplant and CR at day 100 post-alloSCT are favorable

A total of 36 (15.1%) patients underwent alloSCT including nearly 50% of those who had achieved CR1 to prior first-line therapy. Patients with monosomy 7, *TP53* VAF > 25% and *TP53^MH^* at diagnosis were significantly less likely to undergo alloSCT (*P =* .010, *P =* .025, *P =* .050 respectively). Transplanted individuals enjoyed significantly superior median survival (Not reached *vs.* 5.6 mos. for no alloSCT; *P_Log-rank_*< .0001) and OS24 (58.0% *vs.* 5.0% for no alloSCT; P*_fpm_*< .0001). Likewise, among transplanted individuals, ongoing CR at day +100 post-alloSCT was significantly associated with better OS24 (62.7% *vs.* 36.8% for no CR; *P_fpm_ =* .054), in congruence with the COMMAND study demonstrating significant beneficial impact of alloSCT in *TP53^MUT^* MNs.^10^ Early relapse by day+100 was significantly more likely in patients with an *EPI6* signature present (*P =* .002), high EAp53 score (*P =* .018) with a marginal effect for *CUX1* alterations (*P =* .08) at diagnosis.

### *TP53* Risk Score (*TP53*RS) for *TP53^MUT^* Myeloid Neoplasms for patients treated with HMA-based therapies

#### Multivariable model for entire cohort

Based on the predictors relevant in univariable analysis, a multivariable model for the entire cohort was constructed using 5 predictors including antecedent treated myeloid neoplasm, *TP53* VAF, *TP53^sjm^*, monosomies and *EPI6* signature. The models include regular Cox, 45-day landmark Cox, and a competing risk model modeling only leukemia-specific deaths in the competing risk model (See Table 2 and **Figure 5A** for details).

**Figure 5:**
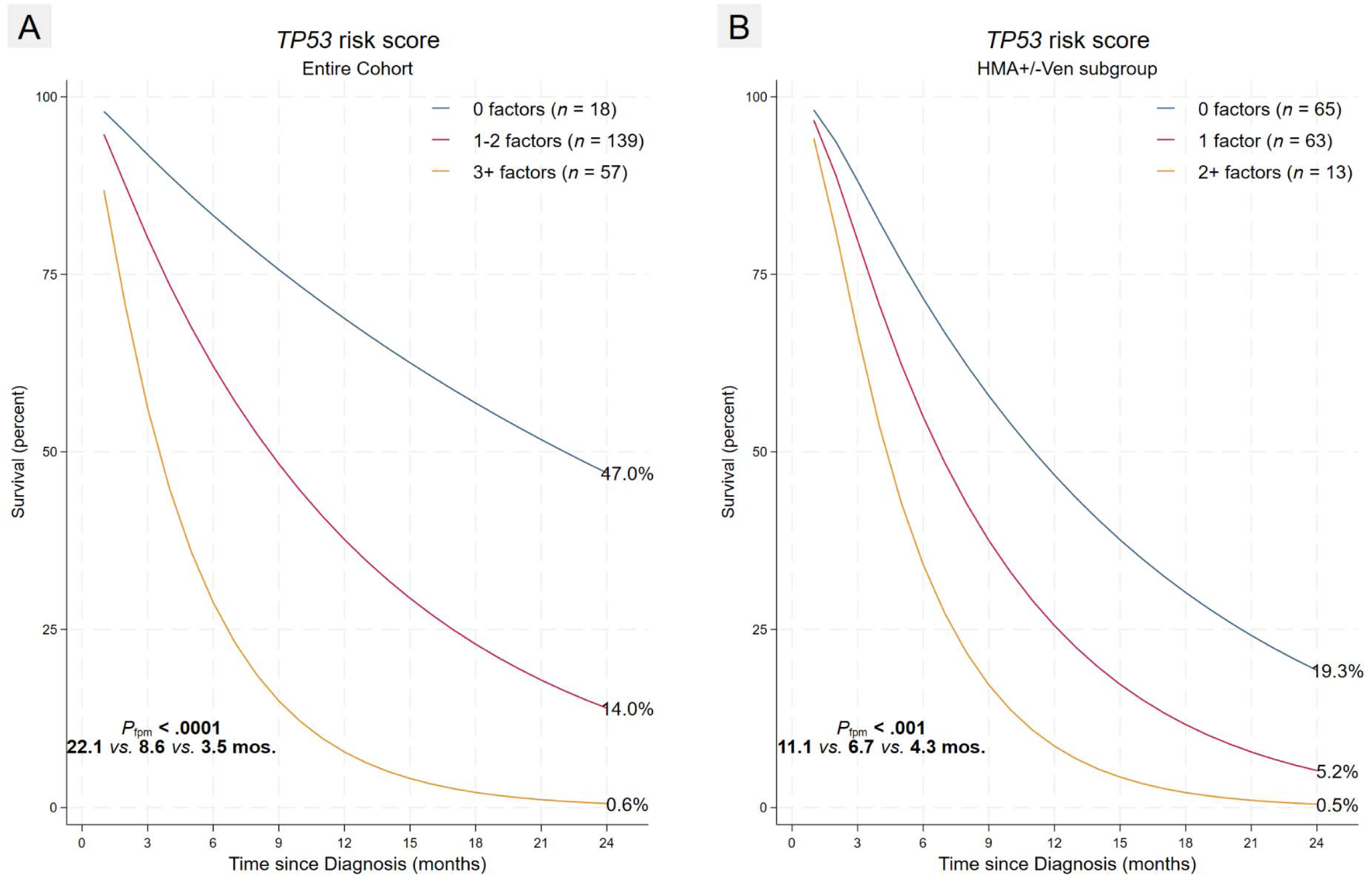
Impact of *TP53* risk score with relevant predictors on OS24 by flexible parametric analysis in entire cohort and HMA subgroup. **A.** Entire cohort analysis utilizing VAF and monosomies in addition to antecedent treated myeloid neoplasm, *TP53^sjm^*and *EPI6* separates three prognostic groups corresponding to risk score cutoffs of 0 factors, 1-2 factors, and 3+ factors. **B.** *TP53* risk score for HMA-treated patients using parsimonious 3 parameter model in sensitivity analysis agnostic to monosomies and VAF information.

**Table 2:** Multivariable (Cox, 45-day landmark Cox and competing risk) models of relevant univariable predictors fitted on the entire cohort incorporating the five relevant covariates adjusted for age at diagnosis.

|  | Cox |  | 45-day landmark Cox |  | Compet. Risk |  |
| --- | --- | --- | --- | --- | --- | --- |
|  | HR | 95% CI | HR | 95% CI | SHR | 95% CI |
| **Age at Dx.** |  |  |  |  |  |  |
| ≤70 yrs | 1.00 |  | 1.00 |  | 1.00 |  |
| >70 yrs | 1.34** | [1.07, 1.68] | 1.45*** | [1.18, 1.78] | 1.23* | [0.96, 1.58] |
| **Anteced. Treated MN** |  |  |  |  |  |  |
| No | 1.00 |  | 1.00 |  | 1.00 |  |
| Yes | 1.72* | [0.93, 3.20] | 1.95* | [0.89, 4.25] | 1.47 | [0.79, 2.71] |
| ***TP53* Splice Mutn.** |  |  |  |  |  |  |
| Absent | 1.00 |  | 1.00 |  | 1.00 |  |
| Present | 1.74* | [0.90, 3.36] | 1.72 | [0.88, 3.34] | 1.79 | [0.81, 3.97] |
| **Highest *TP53* VAF** |  |  |  |  |  |  |
| ≤25% | 1.00 |  | 1.00 |  | 1.00 |  |
| >25% | 1.73*** | [1.35, 2.22] | 1.58*** | [1.31, 1.91] | 1.36** | [1.07, 1.72] |
| **Monosomies** |  |  |  |  |  |  |
| 0-1 monosomy | 1.00 |  | 1.00 |  | 1.00 |  |
| 2+ monosomies | 1.57*** | [1.19, 2.08] | 1.56*** | [1.16, 2.10] | 1.29 | [0.84, 1.97] |
| ***EPI6* signature** |  |  |  |  |  |  |
| Absent *EPI6* | 1.00 |  | 1.00 |  | 1.00 |  |
| Present *EPI6* | 1.86*** | [1.41, 2.45] | 1.80*** | [1.49, 2.16] | 1.43*** | [1.17, 1.75] |

Hazard ratio (HR) for competing risk model denotes Sub-Hazard Ratio
\*\*\* p<.01, \*\* p<.05, \* p<.1

#### Multivariable model and Risk score for HMA-treated subgroup

Since HMA-based therapies are the most predominant frontline choice in this cohort, we developed a separate risk score for this subgroup excluding *TP53* VAF and monosomies (which were relevant only in intensively-treated subgroups as noted earlier)(See Table 3 for details).

**Table 3:** Multivariable (Cox, 45-day landmark Cox and competing risk) subgroup analysis in HMA+/-Ven treated individuals including only three relevant baseline predictors adjusted for age at diagnosis. These three (antecedent treated myeloid neoplasm, *TP53^sjm^*and *EPI6*) were used in development of the *TP53*RS for this group of individuals.

|  | Cox |  | 45-day landmark Cox |  | Compet. Risk |  |
| --- | --- | --- | --- | --- | --- | --- |
|  | HR | 95% CI | HR | 95% CI | SHR | 95% CI |
| **Age at Dx.** |  |  |  |  |  |  |
| ≤70 yrs | 1.00 |  | 1.00 |  | 1.00 |  |
| >70 yrs | 1.37*** | [1.17, 1.61] | 1.44*** | [1.16, 1.80] | 1.28** | [1.01, 1.61] |
| **Anteced. Treated MN** |  |  |  |  |  |  |
| No | 1.00 |  | 1.00 |  | 1.00 |  |
| Yes | 1.74** | [1.12, 2.72] | 2.09** | [1.14, 3.81] | 1.55* | [0.92, 2.59] |
| ***TP53* Splice Mutn.** |  |  |  |  |  |  |
| Absent | 1.00 |  | 1.00 |  | 1.00 |  |
| Present | 2.03** | [1.05, 3.91] | 2.07** | [1.02, 4.22] | 1.96 | [0.80, 4.82] |
| ***EPI6* signature** |  |  |  |  |  |  |
| Absent *EPI6* | 1.00 |  | 1.00 |  | 1.00 |  |
| Present *EPI6* | 1.79*** | [1.45, 2.20] | 1.74*** | [1.50, 2.03] | 1.47*** | [1.23, 1.76] |

Hazard ratio (HR) for competing risk model denotes Sub-Hazard Ratio
\*\*\* p<.01, \*\* p<.05, \* p<.1

This *TP53* risk score (*TP53*RS) for HMA-treated individuals delineated three risk groups comprising intermediate (0 factors, *TP53*RS0), intermediate-poor (1 factor, *TP53*RS1), and poor (2+ factors, *TP53*RS2) risk groups with significantly different median survival (12.6 *vs.* 6.3 *vs.* 4.3 mos.; *P_Log-rank_*< .001) as well as 24-month survival (19.3% *vs.* 5.2% *vs.* 0.5%; P*_fpm_* < .001) respectively (**Figure 5B**). Patients in the poor risk group (2+ factors) were marginally older than 70 years of age (76.9% *vs.* 49.2%*; P =* .06) with inferior frontline response (11.1% *vs.* 31.5% CR1 in 0-1 factors; *P =* .20) with none of these patients receiving an alloSCT.

#### Sensitivity analysis excluding low TP53 VAF patients

Since inclusion of cases with *TP53* VAF < 10% could potentially inflate the impact of the proposed score, we performed an additional sensitivity analysis excluding 17 (7.1%) such individuals. *TP53*RS retained its prognostic value in this last analysis as well (HR = 1.8 [1.3–2.5]; *P <* .001; *N =* 130).

## DISCUSSION

While most prior studies focused on comparing the impact of pretherapy predictors (any *TP53* mutations or *TP53* VAF) in myeloid neoplasms across all risk strata, our cohort is remarkable for being the first to look with just the ELN 2022 high-risk subgroup and identify several novel factors that reliably predict poor response to first-line therapy (monosomal karyotype, multi-hit *TP53* allelic state, *CUX1* deletions) and inferior survival including a novel *EPI6* co-alteration signature including *CUX1*. We specifically focused on developing a model/risk score solely with pre-therapy predictors rather than well-known therapy-related factors (such as CR1 and transplant).

While the proportion of patients treated with chemotherapy was still small in our cohort, it is remarkable that *TP53* allelic state, *TP53* VAF and monosomies were all predictive for survival only in the intensively treated subgroup of patients. While the 25% *TP53* VAF cutoff in our study was admittedly data-driven, it was nevertheless in line with the 23% cutoff proposed by Bahaj and coworkers^22^ as well as the 25% cutoff used by Grob and coworkers^5^ although this latter study was an exclusively chemotherapy-treated, and slightly younger cohort. In comparing the impact of VAF by treatment subgroups, we note as well that *TP53* VAF is relevant only within the intensive chemotherapy treated subgroups, in line with prior studies.^17, 23^ We were however surprised by the higher CR rates in females possibly attributable to less frequent *TP53^MH^*in females. Whether additional fitness-related factors or better compliance in females play a role remains to be seen.

While *TP53* allelic state did not impact survival, it nevertheless remains useful in assessing response to front-line therapy (more so in patients receiving non-intensive therapies; data not shown). Looking at the different *TP53* mutation classes, patients with any *TP53^NMIS^*mutation face significant early mortality. Two prior studies^24, 25^ evaluating MDS demonstrated worse OS with *TP53^NMIS^* mutations compared to *TP53^MIS^* with the latter looking at post-transplant survival. However, an AML study from the German-Austrian AMLSG however did not find any impact on OS based on mutation class (but with a trend towards worse event-free survival with *TP53^NMIS^*) in a cohort treated primarily with intensive chemotherapy.^26^ In our cohort, *TP53* splice junction mutations were associated with the worst outcomes compared to all other mutations. While they occurred much more frequently in men without any impact on CR1 rates, their adverse impact on survival was much more pronounced in women. These findings are congruent with recent demonstration of gender specific differences in the frequencies of AML-associated genetic alterations and less frequent complex karyotype and mutation in genes related to the spliceosome machinery in women (including *U2AF1* and *SRSF2*). ^27^

On the other hand, the predictive relevance of autosomal monosomies is quite remarkable and admittedly somewhat surprising in our high-risk cohort with complex karyotype. While it has long been known that monosomal karyotype is adverse and often enriched in complex karyotype AMLs^4, 28^ with frequent del(17p)^29^, these cohorts comprised a heterogeneous mix of AML patients of all risk groups without specifically focusing on *TP53*-mutated individuals. Considering the high frequency of complex karyotype in our cohort, we restricted the analysis to monosomies only while ignoring structural alterations which were near-universal. Importantly, monosomy 7 was more relevant than monosomy involving chromosomes 5 or 17. We observed that the predictive value of 2+ monosomies for inferior CR1 was restricted only to the subgroup receiving non-intensive therapies underscoring its relevance in an increasingly HMA-treated elderly cohort. However, neither monosomies, nor *TP53* VAF nor allelic state impacted overall survival in the HMA-treated subgroup.

In the context of the *EPI6* signature, *CUX1* emerged as the most pivotal gene. While previous studies^30, 31^ have highlighted the detrimental effects of *CUX1* deletions and mutations in AML, their significance in *TP53^MUT^* MN has not been evaluated. Notably, in our cohort, *CUX1* losses (which were more prevalent than mutations) were observed in 10% of cases. These losses identified a subgroup with an abysmal response rate (0%) to frontline treatment with significantly worse 2-year survival (**Figure 4A**) particularly among female patients. It is important to emphasize that while *CUX1* alterations frequently co-occurred with losses of chromosome 7/7q^32^, isolated losses at this genomic locus were still associated with adverse clinical outcomes, underscoring their independent relevance beyond karyotypically detectable -7/7q. Our observations of *RAS* pathway co-alterations (*CBL*, and *KRAS*) in the *EPI6* signature aligns with the documented activation of this pathway in *CUX1*-altered MNs, as recently demonstrated in the context of 7q alterations in myeloid neoplasms.^33^

We note as well that blast counts are irrelevant once there are more than 9% blasts, affirming that MDS/AML and AML with *TP53^MUT^* are indeed a single biologic entity. That said, establishing a diagnosis of ’morphologic leukemia-free state (MLFS)’ based on blast counts however is particularly problematic in *TP53^MUT^* MN where significant erythroid-predominant leukemic hematopoiesis (often CD34-/strong p53+) is frequent. Post-therapy marrow biopsies frequently show less than 5% CD34-expressing blasts with a significant component of strong p53-expressing erythroid component (frequently corresponding to a mutation) highlighting that neither blast count nor CD34 are good surrogates for pathologic CR or MLFS in *TP53^MUT^* MNs.

To this end, the fast turnaround time of IHC compared to all the other tests (NGS, and karyotype) makes it an attractive surrogate for most if not all non-truncating *TP53* mutations^34^, particularly in elderly AMLs and therapy-related AMLs at diagnosis and after therapy.

Two major limitation of our study include the lack of molecular MRD in response assessment lack of an external validation cohort. To the second point, most published AML and MDS cohort lack public data on alterations (especially losses) at the *CUX1* locus hampering their use as a validation cohort for the *EPI6* signature. Furthermore, previous studies included greater of chemotherapy treated patients limiting their use as a representative validation cohort even if there was available NGS data. Lastly, we were unable to ascertain if some of the observed *TP53* mutations in patients with low VAF merely corresponded to age-related clonal hematopoiesis-associated mutations unrelated to the main clone (see **supplementary figure 6** for analysis of the missense *TP53* substitutions and *TP53* domain relevance in the University of Chicago cohort). Importantly, our work underscores that not all classes of *TP53* mutations are equal given the adverse outcomes with splice junction mutations and early mortality with any *TP53^NMIS^* mutations.

In the real-world setting, all candidate components of the proposed risk score (NGS, FISH, karyotype) are typically available within two weeks of diagnosis. Furthermore, most candidate genes of *EPI6* are common genes included in standard NGS panels, except perhaps *CUX1*, for which we recommend an algorithm in the NGS pipeline for calling *CUX1* loss detection. Among HMA-treated individuals, patients in intermediate/*TP53*RS0 risk group with 0-1 monosomies enjoy a 63% CR1 (*vs.* 16% CR1 with 2+ monosomies; P < .001) are most likely to respond significantly to frontline therapies and have the best chance at alloSCT with durable post-transplant survival. However, most other patients in intermediate-poor/*TP53*RS1 and poor/*TP53*RS2 groups still face early mortality due to comorbidities, active second malignancies or treatment-emergent adverse effects including neutropenic sepsis (especially with venetoclax). As a result, these latter groups derive little to no benefit with existing therapies, highlighting the need for the developing novel agents with better safety and efficacy profiles for these subgroups.

In conclusion, several novel proposed pre-therapy biologic predictors (*CUX1* deletions, allelic state, *TP53* splice junction mutations, *TP53* VAF, autosomal monosomies, and specific co-mutation patterns) as well as antecedent treatment (HMA exposure) for a myeloid neoplasm have differential prognostic utility depending on treatment subgroups (intensive *vs.* HMA-based) and will better inform frontline response and aid in identifying the best candidates for alloSCT in this high-risk, elderly cohort.

## Data Availability

Relevant data can be shared upon reasonable request to corresponding author.

## Acknowledgements

Benjamin L. Ebert, Harvard Medical School, for helpful insights related to biology aspects (CHIP) in email discussions. Esha Patil, undergraduate at University of Illinois, Urbana-Champaign for pre-processing karyotypes prior to parsing with *R::CytoGPS*. Clyde B. Schecter, Nicholas Cox and other members of Statalist for their statistical input (related to data management and analysis) at several time points. Drs. James Vardiman, and Elizabeth Hyjek who were involved in pathologic diagnosis of several earlier cases from University of Chicago included in the study.

## Prior presentation

Portions of these data were presented as an oral presentation as the annual meeting of the United State Canadian Academy of Pathology meeting in 2023 held in New Orlean, LA, USA with another upcoming moderated presentation in the upcoming 2024 USCAP meeting in Baltimore, MD, USA.

## Prior peer review

Peer reviewers of *Blood* for an earlier version of the study submitted to Blood including 207 patients. Most comments/criticisms raised are incorporated/addressed in the current version with a larger cohort size.

## Authorship information

A.K. and A.E.R performed chart review, curated mutations and edited manuscript. P.M.S., and K.K. performed chart review. E.S. performed chart review and collated scripts for figures and manuscript. H.V.S. Visiting pathologist curated data from IARC and EAP53 server. S-Y.Y collated all mutations for lollipop plot generation, B.A., M.S., J.L.P, A.B P. and M.P.M. contributed cases, did chart review and mutation curation of contributed cases. M.T, J.S. & P.Wang curated mutation data and examined pipeline for NGS data. A.L. and C.F. reviewed cytogenetic data. A.P., M.W.D., C.S. & J.E.C, T.K., provided patient care, accrued patients on clinical trials (HMA/Magrolimab, Gilead), designed some analyses, and edited the manuscript. M.E.M reviewed the data related to *CUX1* biology and edited those portions of the manuscript. L.W., E.H., D.A., J.X.C, S.G., S.B.G, and G.V. reviewed pathology data and edited final version of manuscript. O.L. and P.K. analyzed the *TP53* missense mutations in the University of Chicago subset reported in the supplementary data. P.Wanjari. & P.W. analyzed the CNA data. G.V. initiated, designed, supervised study, conducted all statistical analysis, & wrote the manuscript. All authors edited the manuscript.

## REFERENCES

1. Breems DA, Van Putten WL, De Greef GE, et al. Monosomal karyotype in acute myeloid leukemia: a better indicator of poor prognosis than a complex karyotype. J Clin Oncol. 2008;26(29):4791–7.

2. Breems DA, Lowenberg B. Acute myeloid leukemia with monosomal karyotype at the far end of the unfavorable prognostic spectrum. Haematologica. 2011;96(4):491–3.

3. Kayser S, Zucknick M, Dohner K, et al. Monosomal karyotype in adult acute myeloid leukemia: prognostic impact and outcome after different treatment strategies. Blood. 2012;119(2):551–8.

4. Rucker FG, Schlenk RF, Bullinger L, et al. *TP53* alterations in acute myeloid leukemia with complex karyotype correlate with specific copy number alterations, monosomal karyotype, and dismal outcome. Blood. 2012;119(9):2114–21.

5. Grob T, Al Hinai ASA, Sanders MA, et al. Molecular characterization of mutant *TP53* acute myeloid leukemia and high-risk myelodysplastic syndrome. Blood. 2022;139(15):2347–54.

6. Arber DA, Orazi A, Hasserjian RP, et al. International Consensus Classification of Myeloid Neoplasms and Acute Leukemias: integrating morphologic, clinical, and genomic data. Blood. 2022;140(11):1200–28.

7. Weinberg OK, Porwit A, Orazi A, et al. The International Consensus Classification of acute myeloid leukemia. Virchows Arch. 2022.

8. Khoury JD, Solary E, Abla O, et al. The 5th edition of the World Health Organization Classification of Haematolymphoid Tumours: Myeloid and Histiocytic/Dendritic Neoplasms. Leukemia. 2022;36(7):1703–19.

9. Dohner H, Wei AH, Appelbaum FR, et al. Diagnosis and management of AML in adults: 2022 recommendations from an international expert panel on behalf of the ELN. Blood. 2022;140(12):1345–77.

10. Badar T, Atallah E, Shallis R, et al. Survival of *TP53*-mutated acute myeloid leukemia patients receiving allogeneic stem cell transplantation after first induction or salvage therapy: results from the Consortium on Myeloid Malignancies and Neoplastic Diseases (COMMAND). Leukemia. 2023;37(4):799–806.

11. Cluzeau T, Sebert M, Rahme R, et al. Eprenetapopt Plus Azacitidine in *TP53*-Mutated Myelodysplastic Syndromes and Acute Myeloid Leukemia: A Phase II Study by the Groupe Francophone des Myelodysplasies (GFM). J Clin Oncol. 2021;39(14):1575–83.

12. Nawas MT, Kosuri S. Utility or Futility? A Contemporary Approach to Allogeneic Hematopoietic Cell Transplant in *TP53*-Mutated MDS and AML. Blood Adv. 2023.

13. Dohner H, Estey E, Grimwade D, et al. Diagnosis and management of AML in adults: 2017 ELN recommendations from an international expert panel. Blood. 2017;129(4):424–47.

14. Katsonis P, Lichtarge O. A formal perturbation equation between genotype and phenotype determines the Evolutionary Action of protein-coding variations on fitness. Genome Res. 2014;24(12):2050–8.

15. Cox DR. Regression Models and Life-Tables. J R Stat Soc B. 1972;34(2):187-+.

16. Royston P, Parmar MK. Flexible parametric proportional-hazards and proportional-odds models for censored survival data, with application to prognostic modelling and estimation of treatment effects. Stat Med. 2002;21(15):2175–97.

17. Short NJ, Montalban-Bravo G, Hwang H, et al. Prognostic and therapeutic impacts of mutant *TP53* variant allelic frequency in newly diagnosed acute myeloid leukemia. Blood Adv. 2020;4(22):5681–9.

18. Venugopal S, Shoukier M, Konopleva M, et al. Outcomes in patients with newly diagnosed *TP53*-mutated acute myeloid leukemia with or without venetoclax-based therapy. Cancer. 2021;127(19):3541–51.

19. Pollyea DA, Pratz KW, Wei AH, et al. Outcomes in Patients with Poor-Risk Cytogenetics with or without *TP53* Mutations Treated with Venetoclax and Azacitidine. Clin Cancer Res. 2022;28(24):5272–9.

20. Badar T AE, Shallis RM, Litzow, RM. Comparable Survival of Treatment Naïve *TP53* Mutated Acute Myeloid Leukemia Treated with Hypomethylating Agent Compared to Hypomethylating Agent Plus Venetoclax Based Therapy. Annual Meeting & Exposition; Sunday, December 10, 2023; San Diego, CA: American Society of Hematology; 2023.

21. Badar T, Nanaa A, Atallah E, et al. Comparing venetoclax in combination with hypomethylating agents to hypomethylating agent-based therapies for treatment naive *TP53*-mutated acute myeloid leukemia: results from the Consortium on Myeloid Malignancies and Neoplastic Diseases (COMMAND). Blood Cancer J. 2024;14(1):32.

22. Bahaj W, Kewan T, Gurnari C, et al. Novel scheme for defining the clinical implications of *TP53* mutations in myeloid neoplasia. J Hematol Oncol. 2023;16(1):91.

23. Shah MV, Tran ENH, Shah S, et al. *TP53* mutation variant allele frequency of >/=10% is associated with poor prognosis in therapy-related myeloid neoplasms. Blood Cancer J. 2023;13(1):51.

24. Haase D, Stevenson KE, Neuberg D, et al. *TP53* mutation status divides myelodysplastic syndromes with complex karyotypes into distinct prognostic subgroups. Leukemia. 2019;33(7):1747–58.

25. Lindsley RC, Saber W, Mar BG, et al. Prognostic Mutations in Myelodysplastic Syndrome after Stem-Cell Transplantation. N Engl J Med. 2017;376(6):536–47.

26. Boettcher S, Miller PG, Sharma R, et al. A dominant-negative effect drives selection of *TP53* missense mutations in myeloid malignancies. Science. 2019;365(6453):599–604.

27. Ozga M, Nicolet D, Mrozek K, et al. Sex-associated differences in frequencies and prognostic impact of recurrent genetic alterations in adult acute myeloid leukemia (Alliance, AMLCG). Leukemia. 2023.

28. Jang JE, Min YH, Yoon J, et al. Single monosomy as a relatively better survival factor in acute myeloid leukemia patients with monosomal karyotype. Blood Cancer J. 2015;5(10):e358.

29. Gaillard JB, Chiesa J, Reboul D, et al. Monosomal karyotype routinely defines a poor prognosis subgroup in acute myeloid leukemia and is frequently associated with *TP53* deletion. Leuk Lymphoma. 2012;53(2):336–7.

30. Schwartz JR, Ma J, Lamprecht T, et al. The genomic landscape of pediatric myelodysplastic syndromes. Nat Commun. 2017;8(1):1557.

31. Aly M, Ramdzan ZM, Nagata Y, et al. Distinct clinical and biological implications of *CUX1* in myeloid neoplasms. Blood Adv. 2019;3(14):2164–78.

32. Jotte MRM, McNerney ME. The significance of *CUX1* and chromosome 7 in myeloid malignancies. Curr Opin Hematol. 2022;29(2):92–102.

33. An N, Khan S, Imgruet MK, et al. Oncogenic *RAS* promotes leukemic transformation of *CUX1*-deficient cells. Oncogene. 2023;42(12):881–93.

34. Tashakori M, Kadia T, Loghavi S, et al. *TP53* copy number and protein expression inform mutation status across risk categories in acute myeloid leukemia. Blood. 2022;140(1):58–72.

35. Abrams ZB, Tally DG, Abruzzo LV, et al. RCytoGPS: an R package for reading and visualizing cytogenetics data. Bioinformatics. 2021;37(23):4589–90.

36. Zhou X, Edmonson MN, Wilkinson MR, et al. Exploring genomic alteration in pediatric cancer using ProteinPaint. Nat Genet. 2016;48(1):4–6.

