## Supplemental data/figures for "Real World Predictors of Response and 24-month survival in high-grade *TP53*-mutated Myeloid Neoplasms"

### S1: Methods: Supplementary Cytogenetic methods

**Karyotype**: Chromosome analysis (karyotype) was performed on unstimulated culture cells post processing with Conroy's fixative (3:1 methanol:glacial acetic acid). Metaphase chromosomes were stained for analysis following standard methods. Slides were prepared from fixed cultured cells and aged overnight (12-18 hours) in a 50-60°C oven. Slides were treated the next day with 0.1% trypsin (ThermoFisher Scientific, Waltham, MA), followed by treatment with 10% fetal bovine serum in isoton (ThermoFisher Scientific, Waltham, MA). Slides were then treated with a giemsa stain Gurr buffer (ThermoFisher Scientific, Waltham, MA) and rinsed with water (MilliporeSigma, Burlington, MA). G-banded slides were imaged using the CytoVision GSL-120 automated cell imaging system (Leica Biosystems, Wetzler, Germany). Using standard chromosome analysis (karyotype) techniques upwards of 20 metaphase cells were analysed. Results were reported using the appropriate version of the International System for Human Cytogenomic Nomenclature (ISCN, Karger Publishing Company).

**FISH:** Fluorescence in situ hybridization (FISH) using the standard AML and MDS FISH probes was performed on unstimulated cultured cells post processing with Conroy's fixative (3:1 methanol:glacial acetic acid). Hybridization of cells was performed using standard methods. Slides were prepared from fixed cultured cells using the FISH probes specified above and processed with the ThermoBrite (Abbott Molecular, Downer's Grove, IL) using the following hybridization program: 71°C for 2 minutes and 38°C overnight (12-18 hours). Slides were washed the next day in a series of salt solutions: 0.4xSSC (Fisher Scientific, Hampoton, NH) at 40°C for 2 minutes, 0.4xSSC/0.3%TritonX (Promega, Madison, WI) at 75°C for 2 minutes, and 2xSSC/0.1% TritonX at room temperature for 2 minutes. Slides were counterstained with DAPI II (Sigma-Aldrich, St. Louis, MO). Using standard FISH scoring techniques, 200-400 interphase nuclei were manually enumerated for each target using fluorescent microscopy. Cut-off values for positive results were calculated using the beta inverse function, as part of the clinical validation of each FISH probe8 and results were reported using the appropriate version of the International System for Human Cytogenomic Nomenclature (ISCN, Karger Publishing Company).
Considerations for karyotype: Numbers of monosomies and structural alterations were culled from the ISCN by parsing scripts in Stata 18 that identified only unique autosomal monosomies across multiple clones (rather than one abnormal clone) in any given case. Karyotypes with isolated non-clonal abnormalities were included only if these contained recurrent myeloid alterations involving chromosomes 5 and 7.

### S2: Methods: Additional methodology on NGS

Next-generation sequencing (NGS) was performed using different versions of the University of Chicago Medicine OncoPlus (UCM-OncoPlus) panel, a hybrid-capture panel targeting 1005-1213 cancer-associated genes as previously described.[1] Depending on the versions of this assay, variants identified in 119 to168 genes were clinically reported. Briefly, Genomic DNA is isolated from this specimen using the QIAamp DNA Blood Mini Kit (Qiagen). Following extraction, DNA is quantified using the Qubit fluorometric assay (Thermo Fisher Scientific). DNA is subjected to ult*RAS*onic fragmentation and subsequent library preparation using adapter molecules containing patient-specific index sequences (HTP Library Preparation Kit, Kapa Biosystems). After library amplification, quantification and pooling, fragments originating from targeted genomic regions are enriched using a panel of biotinylated oligonucleotides (Roche Nimblegen, or IDT xGen Lockdown probes) supplemented with additional oligonucleotides (xGen Lockdown Probes, IDT). The pooled libraries were sequenced in rapid run mode on a HiSeq 2500 system or NovaSeq 6000 system (Illumina) to produce 2 x 101 bp paired end sequencing reads. Somatic variant calls were inspected using Integrated Genomics Viewer (IGV; Broad Institute, MIT Harvard, Cambridge, MA). Cosmic and ClinVar databases were used as additional tools to interpret detected variants. The hybrid capture probes included coverage for the most common breakpoints seen in *TP53* gene. Data analysis was performed on a HIPAA-compliant high performance computing system (Center for Research Informatics, The University of Chicago) using an in-house developed bioinformatics pipeline. The most recent panel for somatic testing at University of Chicago comprised 168 clinically reportable genes. All genes related to the *EPI6* signature are depicted in bold and tested by at least two centers.

Table of shared genes across centers: Only genes shared at least across two or more centers are depicted here. Genes not clinically reported at University of Chicago are exclude from the list. *EPI6* genes are bolded.

| Gene | N_shared by | Gene | N_shared by | Gene | N_shared by |
|---|---|---|---|---|---|
| ***KRAS*** | 4 | ***U2AF1*** | 4 | *ABL1* | 3 |
| ***CBL*** | 4 | *JAK2* | 4 | *KDM6A* | 3 |
| ***TET2*** | 4 | *IDH1* | 4 | *FBXW7* | 3 |
| *DNMT3A* | 4 | *RUNX1* | 4 | *GNAS* | 3 |
| ***EZH2*** | 4 | *ETV6* | 4 | *SMC1A* | 3 |
| ***CUX1*** | 3 | *KIT* | 4 | *HRAS* | 3 |
| *NF1* | 2 | *WT1* | 4 | *GATA1* | 3 |
| *GATA2* | 4 | *NRAS* | 4 | *NOTCH1* | 3 |
| *CALR* | 4 | *CSF3R* | 4 | *DDX41* | 2 |
| *ASXL1* | 4 | *IDH2* | 4 | *CDKN2A* | 2 |
| *BCOR* | 4 | *SF3B1* | 4 | *PTEN* | 2 |
| *MPL* | 4 | *CEBPA* | 4 | *STAT3* | 2 |
| *TP53* | 4 | *PTPN11* | 4 | *PDGFRA* | 2 |
| *SRSF2* | 4 | *BRAF* | 4 | *STAT5B* | 2 |
| *PHF6* | 4 | *MYD88* | 3 | *RB1* | 2 |
| *NPM1* | 4 | *BCORL1* | 3 | *ATRX* | 2 |
| *FLT3* | 4 | *IKZF1* | 3 | | |
| *ZRSR2* | 4 | *SMC3* | 3 | | |
| *SETBP1* | 4 | *CBLB* | 3 | | |
| *STAG2* | 4 | *RAD21* | 3 | | |

Additionally, we looked for mutations/alterations in specific sets of gene related to certain pathways, including myelodysplasia-related genes[2] (*ASXL1, BCOR, EZH2, RUNX1, SF3B1, SRSF2, STAG2, U2AF1, or ZRSR2*), spliceosome complex genes (*SF3B1, SRSF2, U2AF1 or ZRSR2*).

### S3: Methods: EAp53 score assessment

Given the preponderance of missense mutations in the cohort, we additionally evaluated a previously described computationally derived score to assess the impact of different missense *TP53* mutations called the evolutionary action score for p53.[3] The EAp53 score, as originally designed, ranges from 0-100 with 0 corresponding to wild-type and 100 with higher scores generally correlating with worse outcome. Other solid organ cancers including head and neck cancers[4] as well as lung cancers[5] have been evaluated where the score has demonstrated prognostic utility and more recently in myelodysplastic syndrome where a score of more than 52 predicted for worse outcome.[6] EAp53 scores for all observed missense *TP53* mutations were culled from the EAp53 server at http://mammoth.bcm.tmc.edu/EAp53. In line with the method of Kanagal-Shamanna et al[6], we used the highest VAF for all statistical analyses in patients with more than 1 missense mutation in multi-hit *TP53* cases. We assessed several cutoffs for assessment of EA*p53* scores via a –foreach- loop (within Stata 17) in Cox PH modeling (in the entire dataset and data subsets) and final analysis included patients in the highest quartile of EAp53 scores (>86) *vs.* the lower three quartiles.

### S4: Methods: Germline testing using skin fibroblasts

Samples included germline (cultured skin fibroblasts) or hematopoietic tissue equivalents (peripheral blood, bone marrow, or saliva). Germline testing criteria included personal and family history of HM as well as the National Comprehensive Cancer Network (NCCN) malignancy-specific guidelines for germline testing.[7] Testing for germline variants was performed in a CLIA-certified clinical genomics laboratory using standard clinical protocols. The panel design including variable numbers of genes with fewer genes in patients prior to 2018. Panel-based sequencing was performed at the University of Chicago as described previously.[1] Variants were interpreted per American College of Medical Genetics and Genomics (ACMG)/Association for Molecular Pathology (AMP) guidelines. A small subset of patients had only targeted germline sequencing of either *TP53* (n=7) or *GATA2* (n=1) to exclude Li Fraumeni Syndrome or a germline *GATA2* predisposition. These patients were not included in the final analysis related to germline data.

### S5: Methods: Additional considerations on statistical analysis

Group means, medians, and proportions were examined using the Student's t-test, Mann-Whitney test, or Fisher exact test, respectively. Most continuous covariates such as age were modeled as factor variable using cutoffs noted in the manuscript. Since the cohort was an older cohort, we used an age cutoff of 70 yrs. Patients noted in clinical chart to have been accepted for terminal hospice treatment were marked as 'deceased related to disease' a week after the clinical note date indicating hospice transfer. For the outcome analysis, main and interaction effects models (as relevant) were fitted to calculate hazard ratios with 95% CI while assessing any violations of proportional hazards (P-H) assumptions using scaled Schöenfeld's residuals. All tests were two-sided, and analysis results were considered statistically significant if the two-sided *P* value was less than 0.05. All models consistently supported the assumption of proportional hazards while Cox–Snell residuals indicated good model fit. Flexible parametric P values are depicted as needed when there was borderline or outright violation of PH assumptions. Both non-parametric and parametric model yielded equivalent results.

We additionally performed competing risk analysis designating deaths not directly related to *TP53$^{MUT}$* MN as competing events (such as transplant-related mortality or deaths related to relapse of non-myeloid malignancy in therapy-related myeloid neoplasm patients). Since there was marginally worse outcome for patients older than 70 years, nearly all graphs presented after competing risk (modeling transplant related and non-myeloid neoplasm-related deaths are competing risk) were age-adjusted survival curves.[8] All analyses report 24mo-OS (OS24) and hazard ratios with 95% confidence intervals. A p-value < 0.05 was considered statistically significant without adjustment for multiple testing.

We evaluated the fit of our models by using Harrell's concordance index or Gonen and Heller concordance index for the models generated after Cox PH regression, and Somer's D for the flexible parametric analyses. Additionally, we performed a landmark analysis at 45 days after diagnosis date (since there was a significant fraction of patients with early mortality either related to myeloid neoplasm, infections related to therapy or co-morbidities related to medical conditions or concurrent active second non-hematopoietic cancer) to evaluate the equivalence of the model fit for the multivariate model and avoid potential immortal time bias.[9]

### Supplementary Figures and Tables

**Figure 1: Impact of different *TP53* VAF cutoffs stratified by gender.** VAF cutoffs at 10% (A) & (B) as well as 25% (C) & (D) are prognostic only in males (A & C), most striking with the 10% cutoffs where males have median survival over 50% at 2 years. The difference was less striking with the 25% cutoff as well as 40% cutoff (not shown).

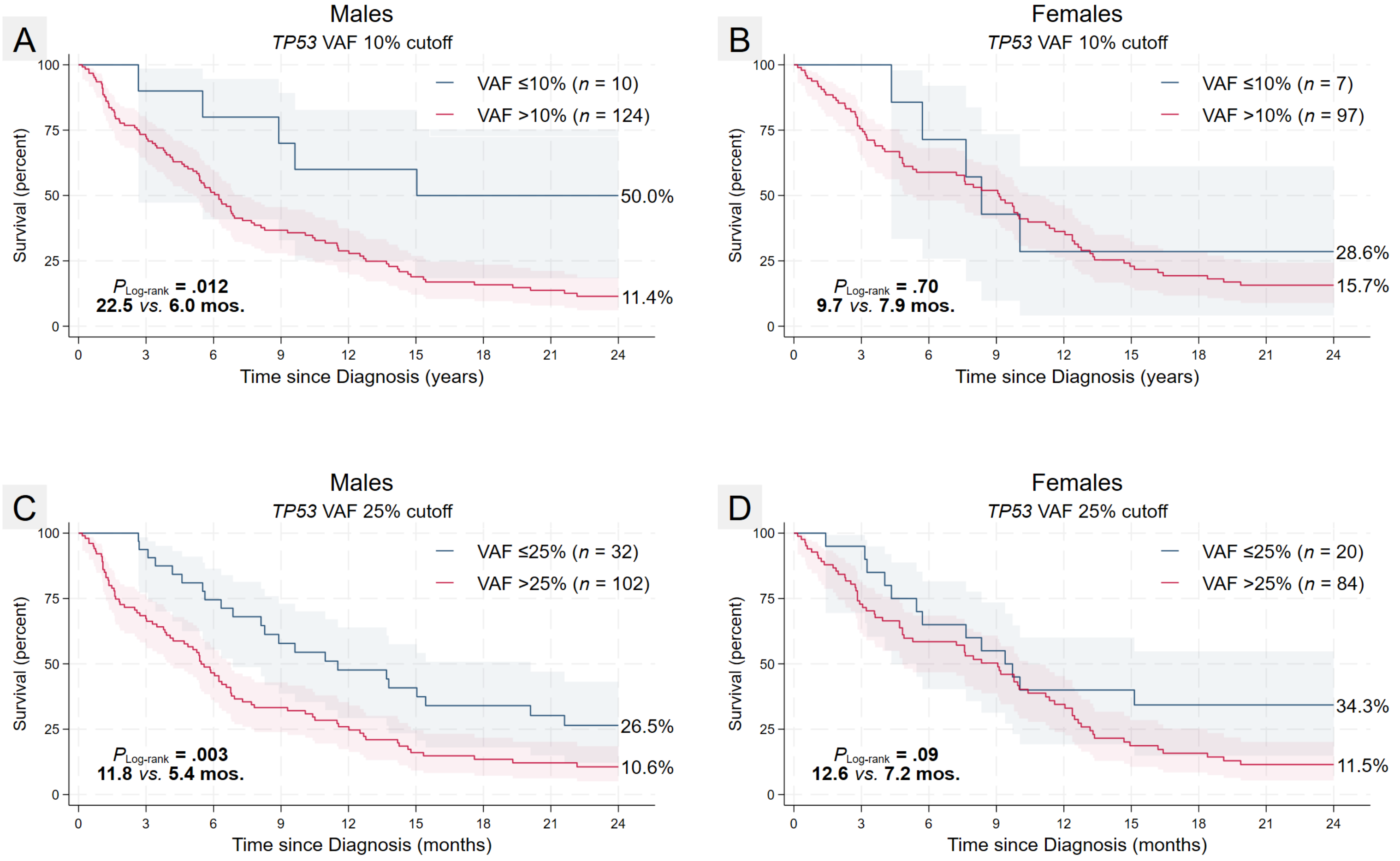

**Figure 2: Receiver Operator Characteristic plots of factors predicting poor frontline treatment response.** Curves were generated after fitting uni-variable age-adjusted logit models for each variable predicting inferior CR1 rates including 2+ monosomies, *TP53*$^{MH}$ allelic state and *EPI6*. All analyses excluded patients on best supportive care for whom response data could not be assessed. In a multi-variable logistic regression model fitted with CR1 as the outcome variable, all three retained their signficance with the most significant contribution from ≥ 2 monosomies while -17/17p was not relevant.

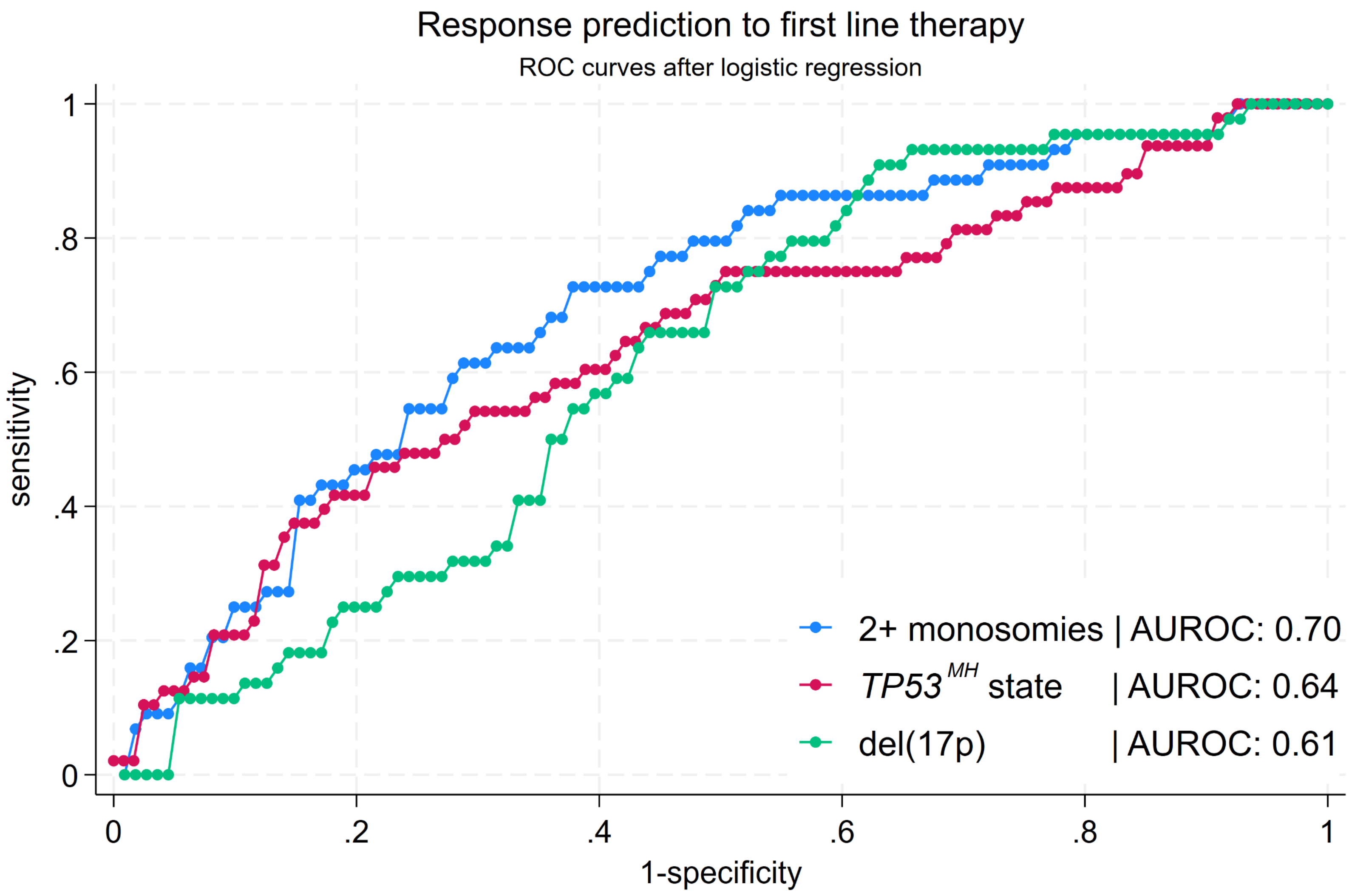

**Figure 3: Outcome stratified by front-line therapy class.** Intensive therapies offered the best outcomes while CPX-351 was associated with the most inferior OS24. In age-adjusted analysis, the beneficial impact of intensive chemotherapy regimens was only marginal compared to the other non-intensive regimens.

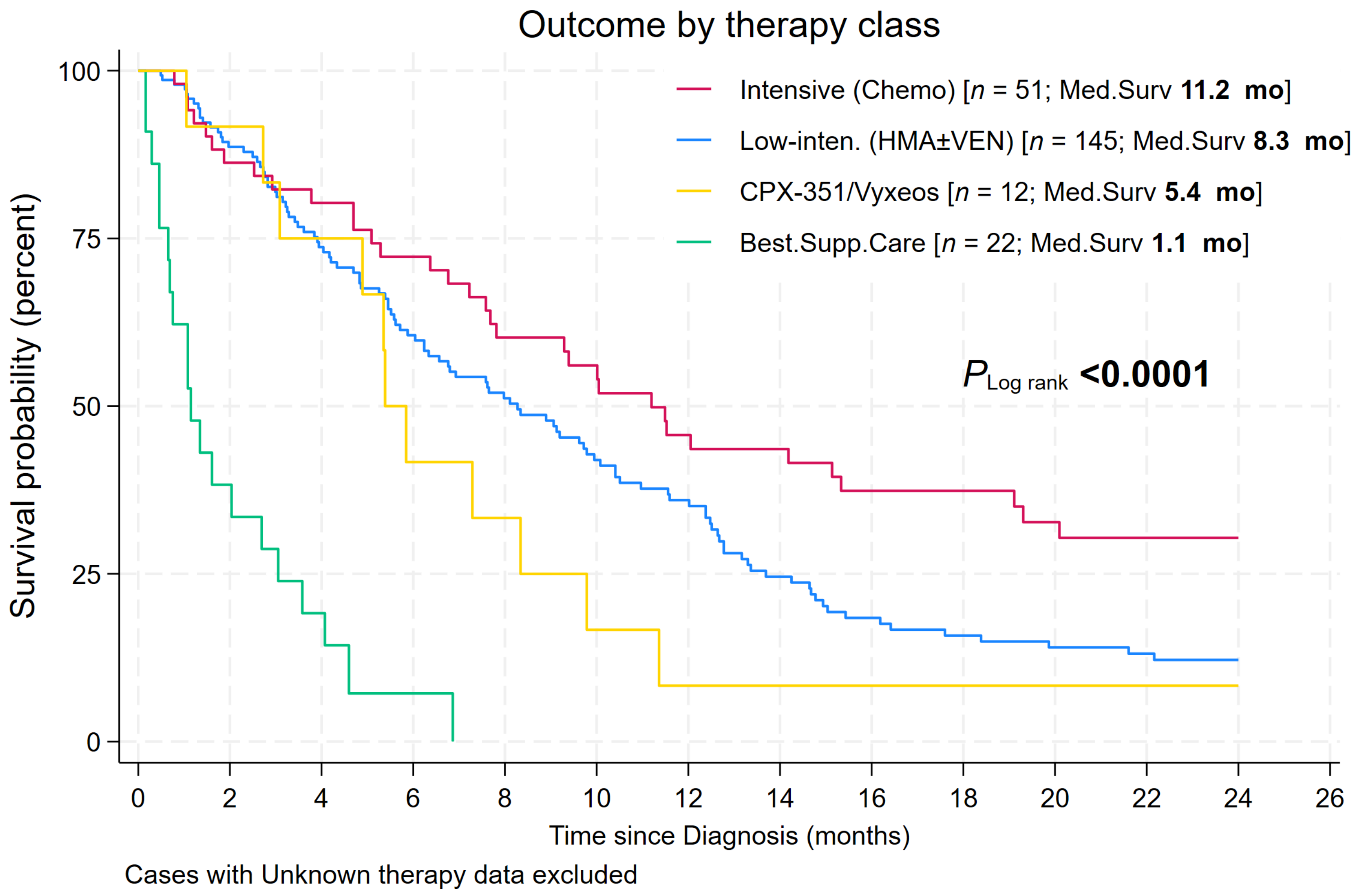

**Figure 4: Highest quartile of Evolutionary Action (EA) p53 (EAp53) score impacts survival in therapy subgroup analysis.** We used a cut-point separating highest quartile (>p75) of EAp53 score (score 86) *vs.* the rest and examined chemotherapy-treated and non-chemotherapy treated subsets, the EA score was relevant only in intensive chemotherapy-treated patients. Furthermore, when we examined the relevance of *TP53* domain (data not shown), patients with L1/S/H2 domain *TP53* mutations had significantly worse outcome (*P* = 0.009 log rank) within the subgroup in the highest quartile of EA score. Based on our observations, the L1/S/H2 motif is very important for tumor development and yet patients have a considerable fraction of mutations with intermediate impact (lower risk). However, when interpreting the survival data, we should keep in mind that the threshold value (highest quartile *vs.* rest) was data driven, so a different threshold may be optimum for a different cancer type. Similarly, we cannot exclude the possibility that a given motif is more important for one cancer type than for another cancer type.

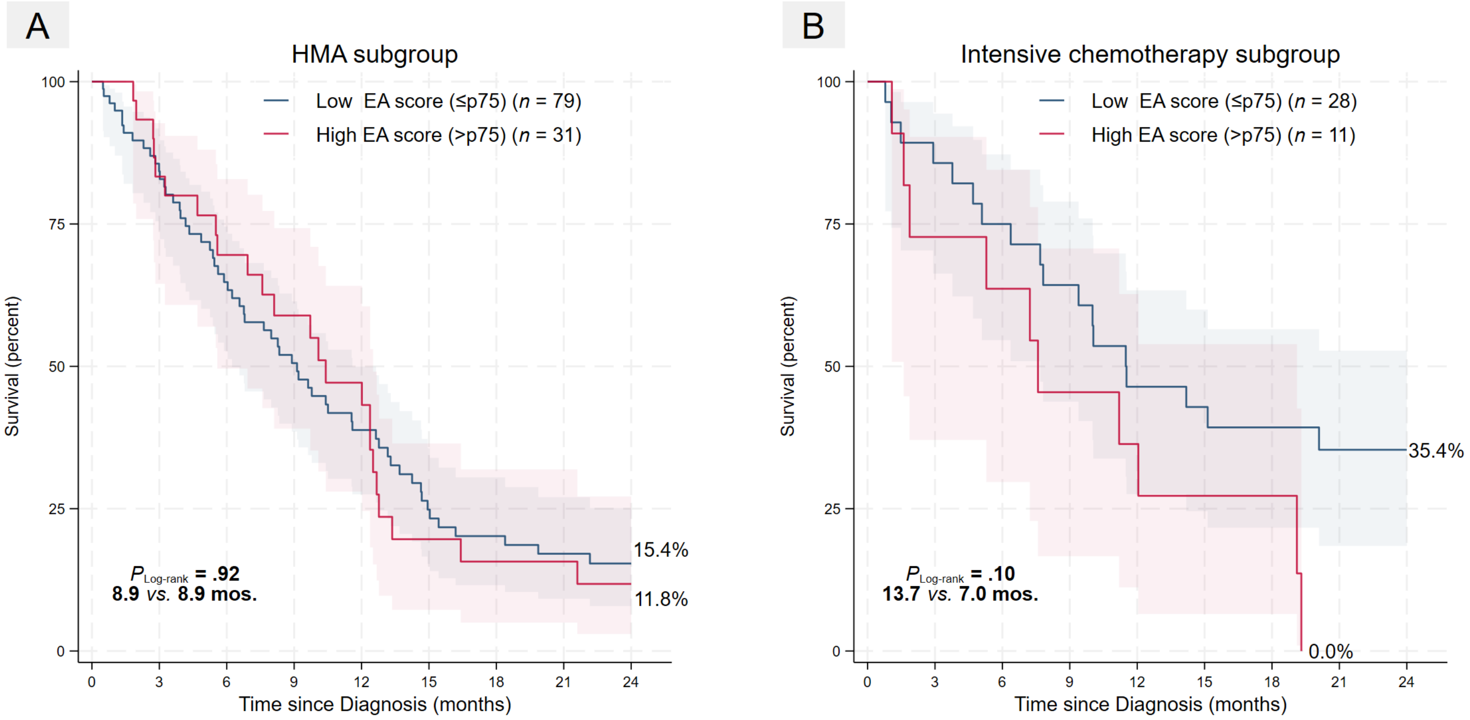

**Figure 5: Forest plots of hazard ratio and 95% CI after uni-variable Cox P-H analysis of pre-therapy covariates by subgroups. A.** *TP53*$^{MH}$ allelic state *vs.* *TP53*$^{SH}$ subgroups and **B.** Therapy class (HMA±venetoclax or intensive chemotherapy groups. While *EPI6* was relevant in both subgroups of allelic state, 2+ monosomies were relevant only in *TP53*$^{SH}$ subset. Looking at relevance of these factors by therapy class, *TP53* VAF > 25% and 2+ monosomies were significantly more relevant in intensive chemo subgroups. Notably *EPI6* was relevant across subsets by allelic state as well as therapy class.

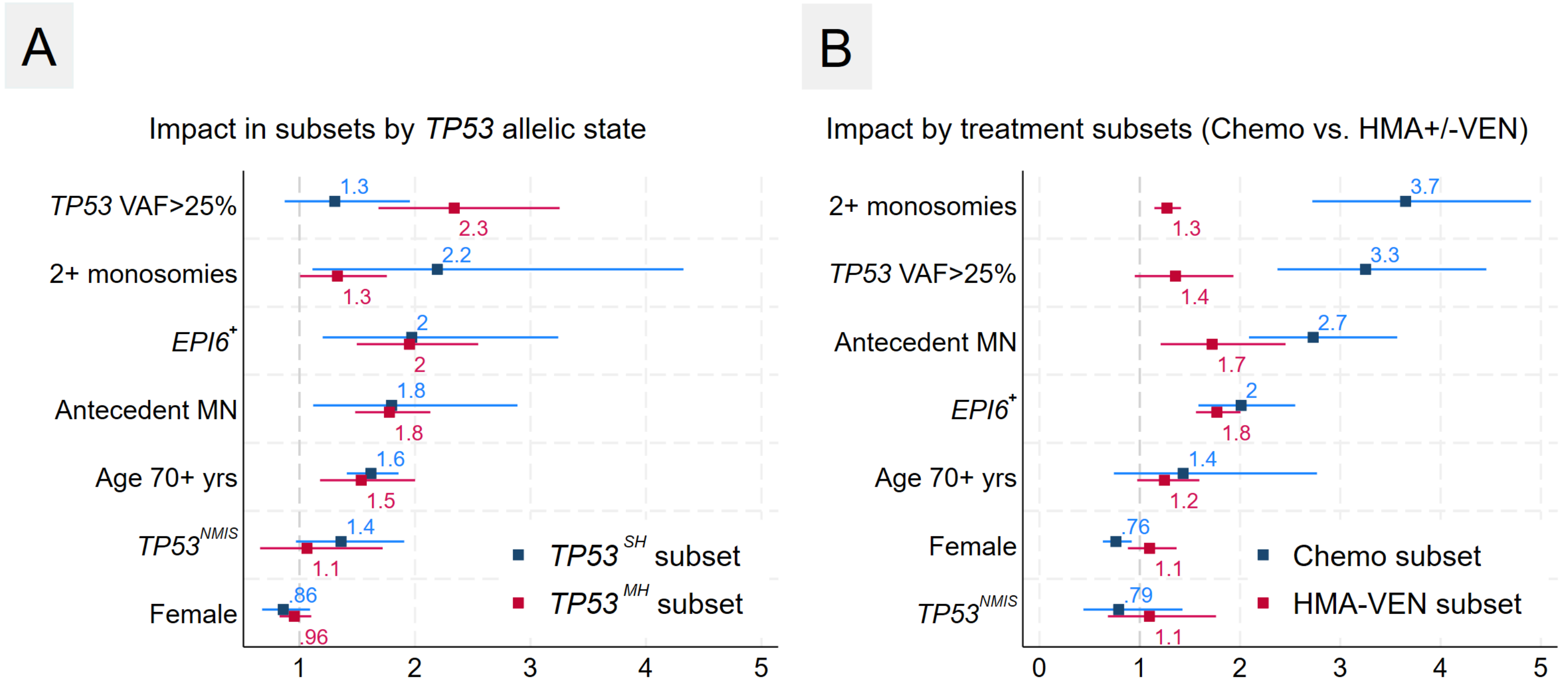

**Figure 6: Functional assessment of *TP53* missense substitutions in the UOC cohort and their relationship to outcome.** Top panel depicts shows the *TP53* structural motifs in our cohort based on the IARC *TP53* database. Middle panel shows the *TP53* DNA Binding Domain structure with the patient mutations shown as spheres for each structural motif. Bottom panel depicts Evolutionary Action (EA) distributions of patient mutations compared to random nucleotide changes for each structural motif.

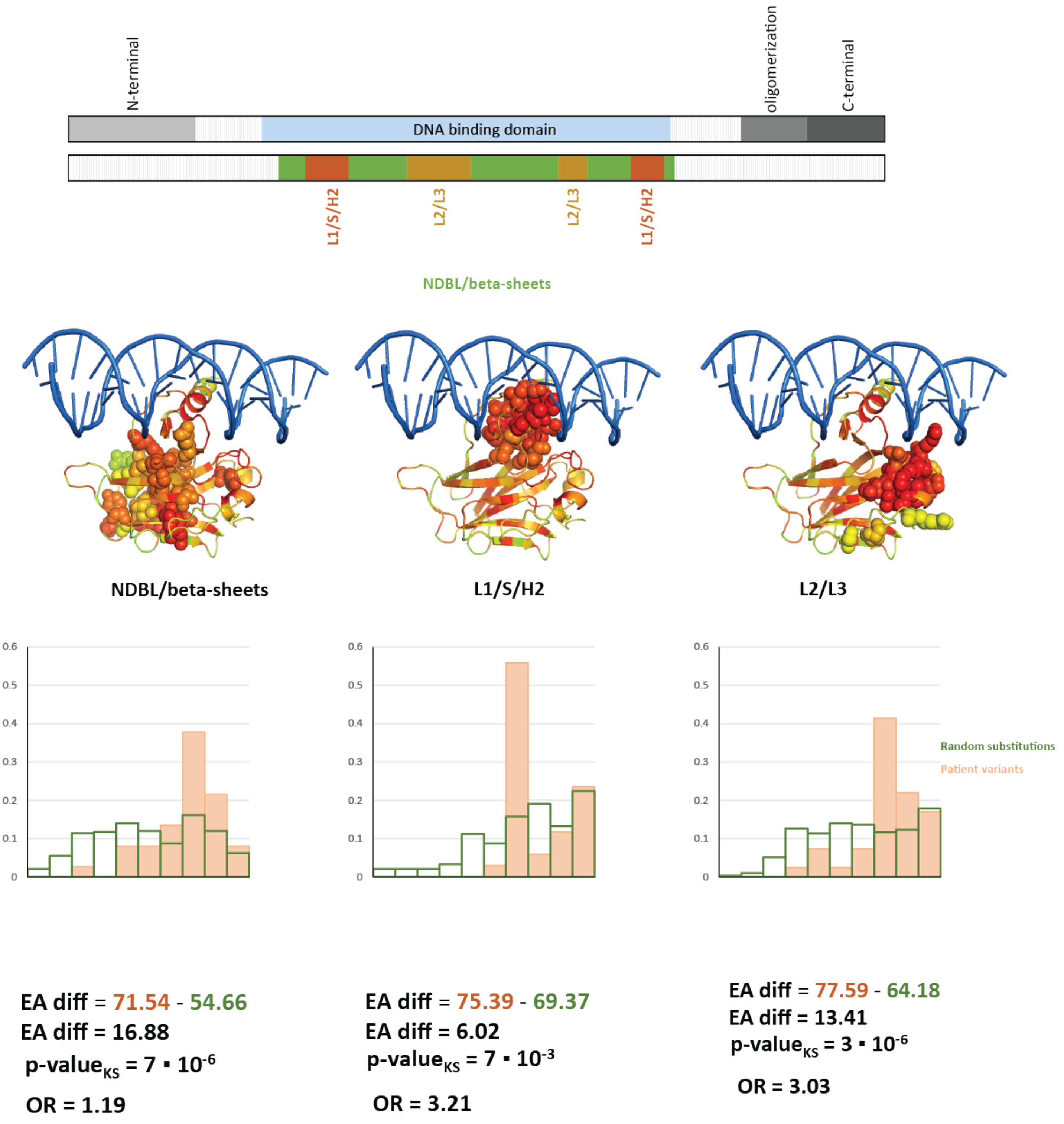

### REFERENCES

1. Kadri S, Long BC, Mujacic I, et al. Clinical Validation of a Next-Generation Sequencing Genomic Oncology Panel via Cross-Platform Benchmarking against Established Amplicon Sequencing Assays. J Mol Diagn. 2017;19(1):43-56.
2. Arber DA, Orazi A, Hasserjian RP, et al. International Consensus Classification of Myeloid Neoplasms and Acute Leukemias: integrating morphologic, clinical, and genomic data. Blood. 2022;140(11):1200-28.
3. Katsonis P, Lichtarge O. A formal perturbation equation between genotype and phenotype determines the Evolutionary Action of protein-coding variations on fitness. Genome Res. 2014;24(12):2050-8.
4. Neskey DM, Osman AA, Ow TJ, et al. Evolutionary Action Score of TP53 Identifies High-Risk Mutations Associated with Decreased Survival and Increased Distant Metastases in Head and Neck Cancer. Cancer Res. 2015;75(7):1527-36.
5. Zhao Y, Han H, Gao Z, et al. Evolutionary Action Score of TP53 Enhances the Prognostic Prediction for Stage I Lung Adenocarcinoma. Semin Thorac Cardiovasc Surg. 2021;33(1):221-9.
6. Kanagal-Shamanna R, Montalban-Bravo G, Katsonis P, et al. Evolutionary action score identifies a subset of TP53 mutated myelodysplastic syndrome with favorable prognosis. Blood Cancer J. 2021;11(3):52.
7. Daly MB, Pilarski R, Yurgelun MB, et al. NCCN Guidelines Insights: Genetic/Familial High-Risk Assessment: Breast, Ovarian, and Pancreatic, Version 1.2020. J Natl Compr Canc Netw. 2020;18(4):380-91.
8. Austin PC, Fine JP. Practical recommendations for reporting Fine-Gray model analyses for competing risk data. Stat Med. 2017;36(27):4391-400.
9. Suissa S. Immortal time bias in observational studies of drug effects. Pharmacoepidemiol Drug Saf. 2007;16(3):241-9.
